# Conventional and unconventional T cell responses contribute to the prediction of clinical outcome and causative bacterial pathogen in sepsis patients

**DOI:** 10.1101/2023.09.13.23295490

**Authors:** Ross J. Burton, Loïc Raffray, Linda M. Moet, Simone M. Cuff, Daniel A. White, Sarah E. Baker, Bernhard Moser, Valerie B. O’Donnell, Peter Ghazal, Matt P. Morgan, Andreas Artemiou, Matthias Eberl

## Abstract

Sepsis is characterised by a dysfunctional host response to infection culminating in life-threatening organ failure that requires complex patient management and rapid intervention. Timely diagnosis of the underlying cause of sepsis is crucial, and identifying those at risk of complications and death is imperative for triaging treatment and resource allocation. Here, we explored the potential of explainable machine learning models to predict mortality and causative pathogen in sepsis patients. By using a modelling pipeline employing multiple feature selection algorithms, we demonstrate the feasibility to identify integrative patterns from clinical parameters, plasma biomarkers and extensive phenotyping of blood immune cells. Whilst no single variable had sufficient predictive power, models that combined five and more features showed a macro area under the curve (AUC) of 0.85 to predict 90 day mortality after sepsis diagnosis, and a macro AUC of 0.86 to discriminate between Gram-positive and Gram-negative bacterial infections. Parameters associated with the cellular immune response contributed the most to models predictive of 90 day mortality, most notably, the proportion of T cells among PBMCs, together with expression of CXCR3 by CD4^+^ T cells and CD25 by mucosal-associated invariant T (MAIT) cells. Frequencies of Vδ2^+^ γδ T cells had the most profound impact on the prediction of Gram-negative infections, alongside other T cell-related variables and total neutrophil count. Overall, our findings highlight the added value of measuring the proportion and activation patterns of conventional and unconventional T cells in the blood of sepsis patients in combination with other immunological, biochemical and clinical parameters.

## INTRODUCTION

Sepsis is a life-threatening syndrome characterised by organ failure arising from a dysfunctional host response to infection. Timely diagnosis of sepsis is crucial, and identifying those at risk of complications and death is imperative for triaging treatment and resource allocation. While severity scores such as Acute Physiology and Chronic Health Evaluation (APACHE) and the Sequential Organ Failure Assessment (SOFA) can be used for audits and to direct care, these tools rely on routinely collected clinical data and observations, and their performance for predicting in-hospital mortality is relatively poor [1, 2]. Most importantly, they do not capture the complex maladaptive immune and metabolic mechanisms that are now known to contribute significantly to sepsis pathology.

The need to identify the cause and target treatment has driven an interest in identifying diagnostic and prognostic biomarkers [3, 4], and in developing algorithms that leverage electronic health records [5, 6, 7]. Whereas single biomarker studies have shown mixed results, the value of multiple biomarkers in combination is increasingly being recognised [4, 8, 9]. In particular, prognostic biomarkers derived from the pathophysiology of sepsis may help guide treatment and monitoring of the disease, the most widely studied ones being C-reactive protein (CRP) and procalcitonin (PCT). However, elevated plasma levels of CRP on admission are not a reliable predictor of mortality [10], and although early levels of plasma PCT differ between survivors and non-survivors, high heterogeneity between study populations puts the general applicability of these findings into question [11, 12]. Other promising biomarkers include pro-adrenomedullin [13], IL-6 [14], lymphopenia [15], neutrophil to lymphocyte ratio [16], and CD64 expression on neutrophils [17], among others.

Most biomarker research in sepsis focuses on diagnosis rather than prognosis, given the importance of early interventions, such as anti-microbial treatments, on survival [18]. In this respect, Gram-negative organisms are often associated with poorer outcomes in first-hit infections [19]. However, the reality of sepsis is that the causative pathogens and the best way to target them are unknown at the time of presentation, and broad-spectrum antibiotics are often administered empirically, with arguments for [20] and against [21] their rapid use. In a significant proportion of patients with sepsis, the causative pathogen is never identified, and there are contradictory findings as to how severity of illness, length of stay and in-hospital mortality compare between culture-negative and culture-positive sepsis [22]. In fact, it is still uncertain whether culture-negative sepsis represents a separate clinical entity, with the possibility of the absence of infection entirely. Earlier identification of the causative pathogen would undoubtedly lead to better targeted therapy and improve antibiotic stewardship [23].

In this respect, multiple molecular methods for pathogen identification have come to market, potentially reducing the time needed to identify the causative pathogen by up to 30 hours [24]. However, many technologies still require a positive blood culture, and sensitivities for direct detection of bacteria in blood by PCR are only modest [25]. Encouragingly, plasma levels of PCT appear to be higher amongst patients with Gram-negative infections compared to Gram-positive infections [26]. Other biomarkers that have shown promise are soluble CD14 (‘presepsin’), which appears to be increased in Gram-negative bacteraemia [27], and the cytokines IL-1β, IL-6, and IL-18, with elevated concentrations in patients with Gram-positive infection [28].

Of particular interest for identifying the aetiology in sepsis are unconventional T cells such as mucosal associated invariant T (MAIT) cells and γδ T cells, which are capable of microbial pattern recognition. MAIT cells are characterised by a semi-invariant T cell antigen receptor with specificity for microbial riboflavin (vitamin B2) derivatives found in fungi and most bacteria except *Streptococcus* spp., *Enterococcus* spp. and *Listeria monocytogenes* [29]. Human Vγ9/Vδ2^+^ γδ T cells respond to the microbial isoprenoid precursor (*E*)-4-hydroxy-3-methyl-but-2-enyl pyrophosphate (HMB-PP), a molecule produced by most Gram-negative and some Gram-positive pathogens, but notably absent from *Streptococcus* spp., *Enterococcus* spp., *Staphylococcus* spp. and fungi [29]. The innate functionality and specificity of these unconventional T cells are likely to contribute to pathogen-specific ‘immune fingerprints’, with proof of concept already shown in patients with acute peritonitis [30].

Studies have only begun to incorporate a combination of clinically available data with novel biomarkers to create predictive models for sepsis [31, 32, 33]. With the advent of multi-omics technology, there is a growing abundance of data, with the promise that a multi-layered approach to phenotyping the immunological response to sepsis may help identify diagnostic and prognostic signatures with direct application to the clinic [34, 35]. This diverse feature space presents the challenge of analysing extensive high-dimensional data from which informative biomarker combinations are to be found, in a task is analogous to feature selection in machine learning [36]. The minimal yet optimal variables are identified to help reduce model complexity, avert overfitting and improve performance. Numerous feature selection methodologies already exist, each with benefits and disadvantages [37]. Since no single machine learning algorithm will be optimal for every task [38, 39], it is advised to search across multiple solutions and make conclusions based on the performance of observed data. Experimenting with multiple methodologies will reduce the risk of overlooking an informative signature or focusing on a single suboptimal solution.

Data-driven pattern recognition with feature selection has successfully identified predictive signatures in the pathogenic cause of peritonitis [30], prognosis and treatment response in traumatic injury [40], prognosis in COVID-19 [41], and vaccine response [42]. The application of model agnostic methods for the measure of feature importance was recently demonstrated for predicting multiple organ dysfunction in paediatric sepsis [43] and the identification of risk factors in COVID-19 [44]. We here set out to combine routine clinical data with immunological profiling and develop machine learning models describing composite biomarker patterns predictive of mortality or underlying cause in sepsis patients, aiming to create valuable tools to prioritise and direct care in a resource-limited environment. Our findings underscore the power of comprehensive models integrating clinical parameters with an analysis of acute phase proteins, cytokines and lipids, and an extensive immune phenotyping of the cellular compartment. While perhaps unsurprisingly no variable on its own was informative enough to help guide clinical decisions, models combining five or more features predicted 90 day mortality after sepsis diagnosis, and discriminated between Gram-positive and Gram-negative infections. A particularly compelling observation from this study was the importance attributed to MAIT cells and Vδ2^+^ γδ T cells, providing *in vivo* evidence for an involvement of unconventional T cell subsets in the early immune response in many microbial infections.

## MATERIALS & METHODS

### Subjects

A total of 77 patients were enrolled between 2018 and 2021 who were over 18 years old with a diagnosis of sepsis according to the Third International Consensus Definitions for Sepsis and Septic Shock (‘Sepsis-3’) [45]. They were cared for in the intensive care unit at the University Hospital of Wales in Cardiff, and were recruited within 36 hours of the presumed onset of infective illness when they already had or would require arterial cannulation as part of standard treatment [46]. Patients were excluded if they were pregnant or breastfeeding, or were females of childbearing age in whom a pregnancy test had not been performed; had severe immune deficiency (*e.g.* diagnosis of AIDS, or treatment with anti-rejection transplant drugs or high dose corticosteroids); had haematologic malignancy or ongoing chemotherapy; had pre-existing severe liver failure; were adjudged by the admitting clinician to be unlikely to survive for the duration of the study period; were admitted post-cardiac arrest; or had an underlying impairment of higher function that would make it impossible for informed consent to be given upon recovery (*e.g.* severe learning disability).

### Blood samples and clinical records

Whole blood was obtained within the first 36 hours of sepsis from patients with a SOFA score >2 and a suspected infection; cell-free plasma was obtained and frozen within the first hour of sample collection. Mortality rates of 22.1% and 27.3% after 30 and 90 days after sepsis diagnosis, respectively (Supplementary Tables S1 and S2), were in line with a recent multi-centre prevalence study of sepsis in Wales [47]. 52 patients (67.5% of the cohort) had a microbiologically confirmed infection. Three patients had a mixed culture result with undefined causative pathogen, two had influenza A with no bacterial isolates, and one had candidiasis; these patients were excluded from pathogen-specific analyses. No significant differences in patient demographics, severity scores, therapeutic interventions or mortality were observed between patients with Gram-negative, Gram-positive and culture-negative sepsis (Supplementary Table S3). Routine clinical data such as full blood count, liver profile and blood gas data were recorded; variables captured for fewer than five patients were removed from the subsequent analysis, leaving 63 routinely collected variables (Supplementary Table S4).

### Flow cytometry

Neutrophils and monocytes were stained in freshly collected whole blood after red blood cell lysis; T cells were stained after Ficoll-Paque PLUS (Fisher Scientific) separation of peripheral blood mononuclear cells. Cells were acquired on a 16-colour BD LSR Fortessa flow cytometer (BD Biosciences), using the fluorescently labelled monoclonal antibodies listed in Supplementary Table S5. Live single cells were gated based on side and forward scatter area/height and exclusion of live/dead staining (fixable Aqua; Invitrogen). Exclusion of monocytes and B cells in T cell panels was ensured by using anti-CD14 and anti-CD19 in the live/dead staining channel. When identifying monocytes and neutrophils in whole blood, T cells were excluded by their appearance in side and forward scatter area/height, and B cells were excluded by using anti-CD19 in the live/dead staining channel. The flow cytometer was calibrated using BD FACSDiva CS&T research beads (BD Biosciences) prior to acquisition. Compensation for spectral overlap was accounted for using BD CompBeads (BD Biosciences) and a spillover matrix generated in the FACSDiva software. Compensation was checked for errors using the FlowJo software (TreeStar) prior to analysis.

The Harmony algorithm was applied to all cytometry data using CytoPy version 2.0 to align samples whilst reducing the risk of losing biological information [48], and geometric median clustering with weighted voting (GeoWaVe) was performed on batch-corrected data for T cells, monocytes and neutrophils as described before [49]. Ensembles were informed using multiple clustering algorithms popular for analysing cytometry data, providing diverse input for ensembles and preventing biased analysis driven by a single method, including FlowSOM, Phenograph, and SPADE algorithms as well as K-Means and FlowSOM clustering of PHATE embeddings.

### Soluble proteins in patient plasma

Cytokines and chemokines were quantified in two batches using Luminex standard sensitivity magplex assays based on the xMAP (multi-analyte profiling) technology, according to the manufacturer’s instructions. Data were acquired on a Luminex 200 compact analyser. TNF-α (eBioscience), IFN-γ (eBioscience) and IL-6 (R&D Systems) were measured in a single batch by ELISA. Concentrations were obtained by fitting a standard curve using a five-parameter logistic fit using the generalised Hill equation, and data quality was assessed by observing the coefficient-of-variation and standard recovery. Where fewer than three observations fell within the standard range, the majority of observations had a CV greater than 50%, or the standard recovery was outside a range of 75-125%, analytes were deemed of poor quality and excluded from subsequent analysis. Batch effect was addressed with post-hoc correction and data alignment; data were log_2_ transformed and values replaced with a *z* score. 95% confidence intervals for odds ratios were approximated as previously described by Tenny and Hoffman [50].

### Free fatty acid and acyl carnitine analysis

Concentrations of free fatty acids and acyl carnitines specified in Supplementary Table S6 were determined by liquid chromatography-tandem mass spectrometry (LC-MS/MS). Lipids were extracted from thawed cell-free plasma in randomised batches by mixing 10 μl plasma with 240 μl methanol and 10 µl of a mixture of deuterated internal lipid standards (Supplementary Table S7) for a description of internal lipid standards and concentrations). Samples were sonicated in iced water for 1 min, vortexed at 1,400 rpm for 10 min at 4 °C, and then centrifuged at 18,000 × *g* for 10 min. 100 μl supernatant was derivatised using 3-nitrophenylhydrazine [51], by adding 50 µl 200 mM 3-nitrophenyl-hydrazinein (50/50 methanol/H_2_O) and 50 µl 120 mM N-ethyl-N′-(3-dimethylaminopropyl) carbodiimide hydrochloride and 6% pyridine (50/50 methanol/H_2_O). Samples were then vortexed and incubated for 30 min at 40°C. Excess derivatisation reagents were quenched by the addition of 0.5 % formic acid (100 µl; 75/25 methanol/H_2_O) and incubation at 40°C for 30 min. Samples were aliquoted into HPLC vials for LC-MS/MS. For every sample batch, five blanks of HPLC water were extracted and derivatised to account for background levels of lipids.

LC-MS/MS analyses were performed using scheduled Multiple Reaction Monitoring (MRM) mode on a Nexera liquid chromatography system (Shimadzu) coupled to a QTRAP 4000 mass spectrometer with an ESI source (AB Sciex). Lipid separation was achieved using a Kinetex Polar C18 reverse phase column (100 Å, 100 × 2.1 mm, 2.6 µm particle size; Phenomenex) and gradient elution of two mobile phases (A: 100 % water + 0.1 % formic acid, phase B: 100 % methanol + 0.1 % formic acid). The flow rate was 0.2 ml/min and injection volume was 2 µl; column temperature was 50°C. MS settings (declustering potential and collision energy) were optimised for individual lipids using standards. Fatty acids and acyl carnitines were detected in negative ion and positive ion mode, respectively. All MRMs used are provided in Supplementary Table S7. Calibration curves and a QC sample of pooled sample aliquots were quantified alongside sample analyses. QC samples were run after every ten measurements to monitor MS performance. Peak areas were integrated using MultiQuantsoftware (AB Sciex) and concentrations calculated based on internal standard response. For lipids with an exact corresponding internal standard, concentrations were calculated directly based on the lipid to internal standard ratio and internal standard concentration, while ensuring that all peaks were within the linear dynamic range of the instrument. For other lipids, concentrations were derived from comparisons with an external calibration curve of the ratio of the specific lipid standard to a deuterated internal standard. Limit of quantitation used signal:noise of >5:1 and at least 5 data points per peak. All further analysis was conducted using R and GraphPad Prism version 8.4.3, and MetaboAnalyst version 4.0 and higher (metaboanalyst.ca).

### Data processing

A MongoDB database was populated with clinical parameters, soluble biomarker measurements and summary statistics of flow cytometry data. Data were combined into a table of 267 features (Supplementary Table S6), which could be broadly categorised into physiology, interventions, point of care testing, clinical laboratory results, protein biomarkers, lipids, proportions of immune cell populations and mean fluorescence intensities of surface markers on immune cells. Many clinical variables demonstrated class imbalance, with the majority class representing more than 70% of patients for each target. Features that were obviously redundant were removed, *e.g.* where information was duplicated or values were equal for all patients. A combination of experimental errors, issues of sample integrity and the sporadic nature of clinical data collection all contributed to missing values (Supplementary Fig. S1). Features that could not be assumed to be missing at random, such as clinical laboratory measurements that were obtained for only a few individuals, were excluded from further analysis. The remaining missing values were imputed using the MissRanger algorithm [52]. Imputation resulted in acceptable out-of-bag (OOB) for both continuous and categorical features, with a median normalised mean root squared error (NMRSE) of 0.68 and <20% categorical features misclassified. Features with an NMRSE >1.0 or >40% missing data were removed from further analysis. Multicollinear features were identified and either replaced with an estimated latent variable, or the variables with the greatest mutual information with the targetvariables were retained (Supplementary Fig. S2).

### Feature selection

After removal of redundant and highly correlated features, 150 features remained. Of these, a set had to be chosen small enough to reduce the risk of overfitting, improve classification accuracy, and ensure models could easily be interpreted. Feature selection algorithms comprised filter and wrapper techniques popular in the biostatistics literature: univariate selection with permutation testing [53], ReliefF [54], Minimum Redundancy-Maximum Relevance (MRMR) [55], Boruta [56], and recursive feature elimination with Support Vector Machines (SVMs) [57]. After running each algorithm for all target variables, the top ten features from each algorithm were chosen to provide five independent feature sets for each target. Downstream analysis included validation of models on equivalent complete case data, and constraining the number of features increases the amount of data available during complete case analysis. Limiting the feature space to a maximum of ten parameters ensured at least five observations were available per feature and made interpretation of model decisions easier. Classifiers were generated for the top three to ten features and compared by classification performance. Overlap between feature selection methods was measured using the pairwise Jaccard index and visualised as heatmap for each target (Supplementary Fig. S3).

### Modelling pipeline for selecting, comparing and inspecting classification algorithms

A 20% subset of the total cohort was randomly selected prior to model and feature selection and kept for independent evaluation of model performance to avoid inflated accuracy due to overfitting (Supplementary Fig. S4A). Eight groups of classifiers (referred to as ‘classifier families’ from here onward) were drawn upon for the task of binary classification: Logistic regression, SVMs with linear or non-linear kernel, Naive Bayes, K-Nearest Neighbours (KNN), Random Forest, Extra Random Forest, and Extreme Gradient Boosting (XGBoost) (Supplementary Fig. S4B). All classifiers were implemented using the Scikit-Learn library [58]. A grid search strategy was employed to tune optimal hyperparameters.

Hyperparameters included L1 and L2 regularisation of varying strengths, polynomial and radial basis function kernels for non-linear SVMs (with multiple degrees for the former and a range of γ for the latter), different distance metrics and number of nearest neighbours for KNN, and multiple hyperparameters for ensembles of tree-based learners controlling parameters such as the depth of trees, number of splits, number of features, and sampling methods. This approach resulted in 216 classification models from a diverse range of classifier families, an approach supported by the aptly named “no free lunch theorem” [38], which suggests that no single algorithm can be optimal for all problems.

Each model was trained on five independent feature sets (Supplementary Fig. S4C), chosen to increase diversity when exploring the feature space without significantly impacting computational requirements. As the optimal number of features might be less than the top ten ranked features presented by each feature selection algorithm, each classifier was trained on the top three through to the top ten features, iteratively. For each classifier, eight models were generated for each feature selection algorithm, totalling 40 models across all possible feature sets and 8,880 models trained across all possible classifiers, repeated for each target. Leave-one-out cross-validation (LOOCV) and 5-fold cross-validation (5-fold CV) were used to select the optimal model before validation on independent hold-out data (Supplementary Fig. S4D). Within each classifier family, the model and feature set combined with the highest LOOCV F1 score was chosen for evaluation and model inspection (Supplementary Fig. S4E).

Cross-validation and holdout performance for the optimal model from each classifier family were compared using ROC curves, balanced accuracy, macro F1 score and macro AUC score. Models were first compared by 5-fold CV balanced accuracy using the non-parametric Friedman test and Nemenyi post-hoc testing [59]. After selecting a model from each classifier family using cross-validation, their performance was validated on independent holdout data, and the top performing models were inspected using SHapely Additive exPlanations (SHAP) [60].

## RESULTS

### Insights from routine clinical data in sepsis patients

To identify possible predictors of outcomes and the underlying cause of the infection in patients presenting with sepsis, we first examined all routinely collected clinical data available within the first 36 hours. While all patients had elevated blood levels of CRP, there were no significant differences between survivors and non-survivors, or between culture-positive and culture-negative patients (Supplementary Fig. S5). However, CRP was higher in Gram-positive infections compared to Gram-negative infections. Arterial lactate [61] was increased in those who died within 90 days but was only different when considering the sample closest to enrolment time and not the average within the 48-hour window (Supplementary Fig. S6). As such, CRP and lactate were only of limited utility as biomarkers to make clinically relevant predictions. The same held true for virtually all other clinical variables (data not shown). In fact, the only routinely collected parameter that showed a significant difference between survivors and non-survivors was the fraction of inspired oxygen (FiO2) value taken closest to the diagnosis of sepsis, with increased levels amongst non-survivors compared to survivors, corroborating findings by others [62]. No other biomarker demonstrated a significant difference relative to mortality, nor to the nature of the causative pathogen (Supplementary Fig. S7).

### Soluble immune mediators in patient plasma

To gain further insight into inflammatory responses in sepsis patients, a large panel of soluble mediators in plasma were quantified (Supplementary Fig. S8). Levels of the chemokine CXCL10 were decreased in non-survivors at 30 days, while IL-15 was increased in non-survivors. However, over 80% of patients had IL-15 levels below the detection limit, and the trends seen for CXCL10 and IL-15 were diminished when observing 90 day mortality. FLT3L levels were moderately increased in culture-positive sepsis patients compared to those without a confirmed infection, and ferritin was decreased in patients with Gram-negative compared to Gram-positive infections. No other plasma analytes were significantly different between any of the groups. An additional analysis was performed using the detection limits as thresholds to create binary variables (Supplementary Fig. S9). Although elevated concentrations of IL-6, IL-15 and OSM were associated with higher 30 day mortality, once accounting for multiple comparisons none of the analytes had a statistically significant odds ratio.

### Immune phenotypes in sepsis patients correlate with clinical outcomes

Given the poor predictive performance of routine clinical data and soluble biomarkers we next analysed the cellular compartment by flow cytometry. The first observation was a significant reduction in T cells as percentage of PBMCs in non-survivors compared to survivors at 30 days (Fig. 1A) and 90 days (Fig. 1B), whereas percentages of monocytes and neutrophils amongst white blood cells were not different between the groups. However, and in line with the importance of HLA-DR expression on circulating monocytes as predictor of immunosuppression and poor outcomes in sepsis [63], monocyte HLA-DR expression was lower in non-survivors compared to survivors at both 30 and 90 days (Fig. 2). A trend was also visible for the cell adhesion molecule CD62L, with increased expression on monocytes in non-survivors.

**Figure 1.**
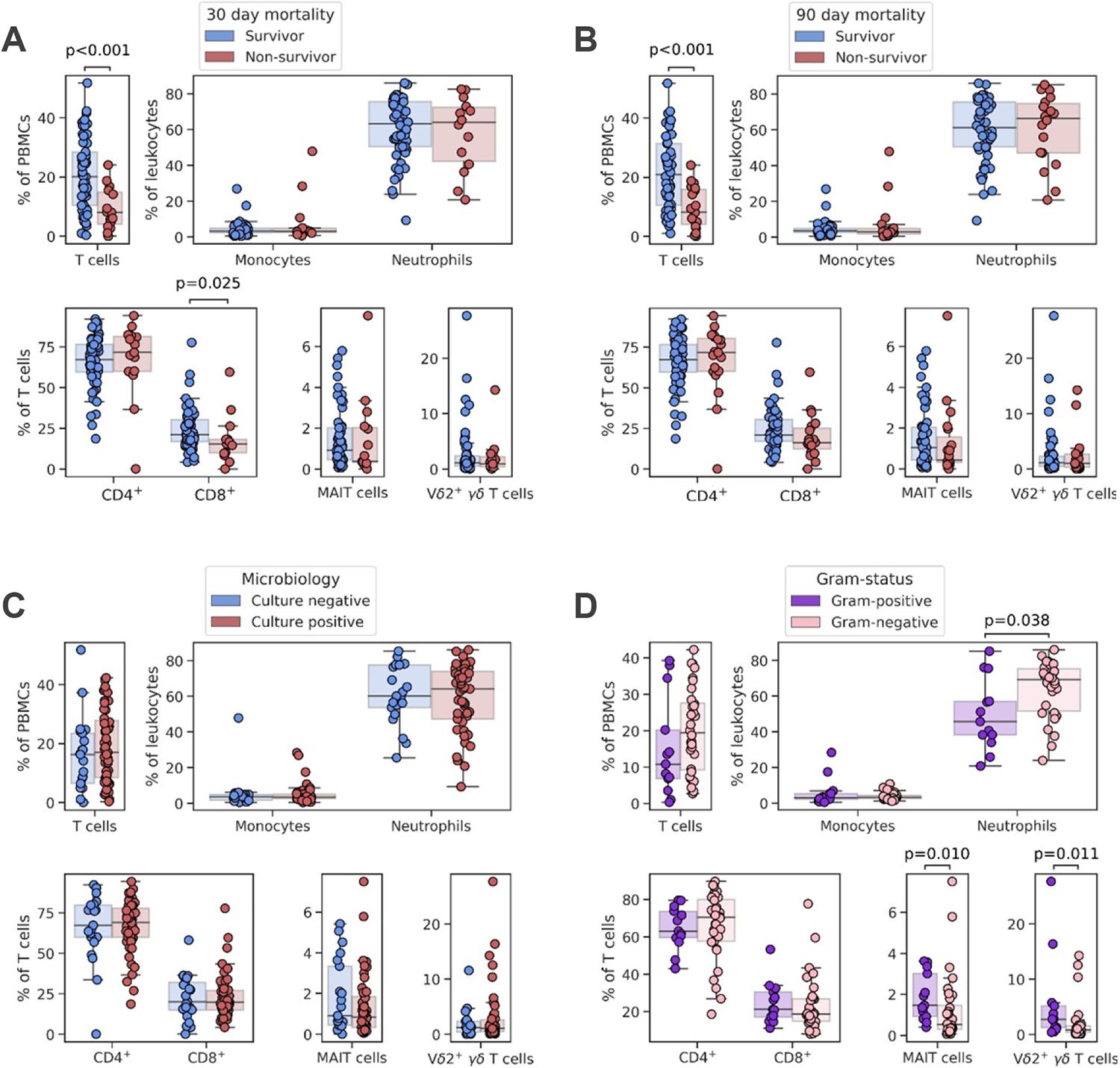
Proportion of T cells, monocytes and neutrophils, and conventional and unconventional T cell subsets in patients after sepsis diagnosis. Comparisons shown are between (*A*) survivors (blue) and non-survivors (red) 30 days after sepsis diagnosis; (*B*) survivors (blue) and non-survivors (red) 90 days after sepsis diagnosis; (*C*) those without (blue) and with (red) a microbiologically confirmed infection; and (*D*) those with a Gram-positive (purple) and Gram-negative (pink) infection, amongst those with a positive bacterial culture. *p* values were generated using two-tailed Mann-Whitney U tests with Bonferroni-Holm corrections for multiple comparisons.

**Figure 2.**
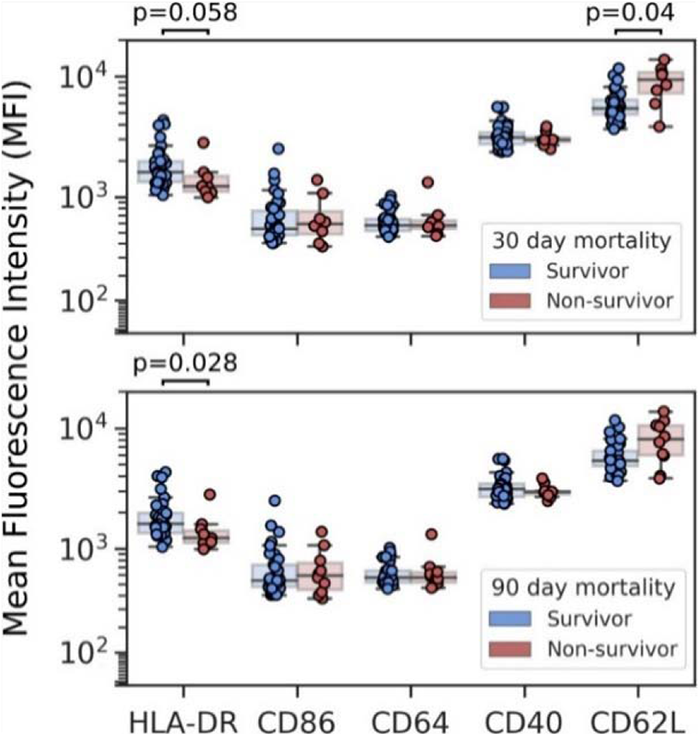
Mean fluorescence intensity (MFI) of HLA-DR, CD86, CD46, CD40 and CD62L on circulating monocytes in sepsis patients. Comparisons between survivors and non-survivors 30 (top) and 90 (bottom) days following a diagnosis of sepsis are shown. *p* values were generated using two-tailed Mann-Whitney U tests with Bonferroni-Holm corrections for multiple comparisons.

Within the T cell compartment (Supplementary Fig. S10), CD8^+^ T cells tended to be reduced in non-survivors, to a greater extent when predicting mortality within 30 days than 90 days. Notably, T cells, monocytes and neutrophils were comparable between those with and without a microbiologically confirmed infection (Fig. 1C). In contrast, there was a significant increase in the proportion of neutrophils amongst white blood cells and a significant decrease in MAIT and Vδ2^+^ γδ T cells as percentage of T cells in patients with Gram-negative infections, compared to Gram-positive sepsis (Fig. 1D). Of note, expression of the activation marker CD69 (and to a lesser extent, of CD25) on MAIT cells and Vδ2^+^ T cells appeared to differentiate Gram-positive from Gram-negative infections, pointing toward a role of these cells in the immune response to some pathogens but not others (Fig. 3).

**Figure 3.**
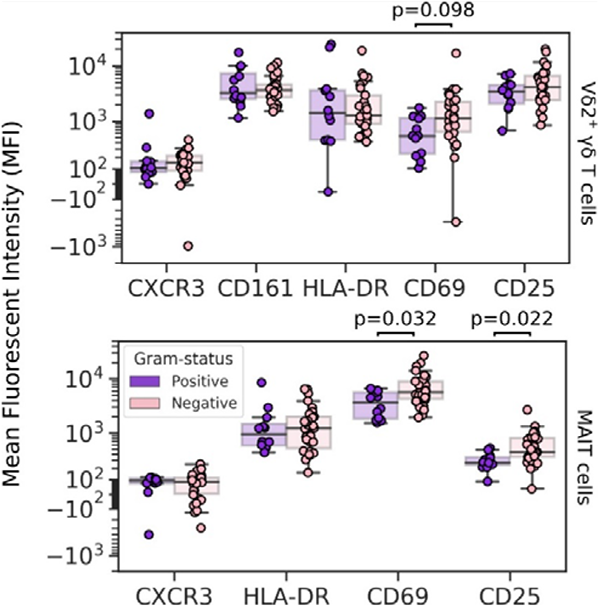
Mean fluorescence intensity (MFI) of HLA-DR, CD86, CD46, CD40 and CD62L on MAIT cells, with comparisons between sepsis patients with a Gram-positive versus a Gram-negative infection. *p* values were generated using two-tailed Mann-Whitney U tests with Bonferroni-Holm corrections for multiple comparisons.

### A T cell dominated immune signature predicts mortality at 90 days after diagnosis of sepsis

We next combined all available information to generate predictive models. There was no significant difference between the optimal models chosen for each classifier family when comparing 5-fold CV accuracy and F1 score for the prediction of 30 day mortality (Supplementary Fig. S11). Logistic regression and SVMs showed promise at first, with excellent LOOCV ROC and comparable performance between training and testing data within the model and feature selection process. However, none of the models generalised well when exposed to holdout data (Table 1). Prediction of 90 day mortality was more reliable, with good LOOCV and 5-fold CV performances across all classifiers (Supplementary Fig. S12). While more complex models such as KNN and ensembles of tree-based learners exhibited more over-fitting compared to the simpler logistic regression and linear SVM, the Extra Random Forest model showed superior accuracy, F1 score and AUC scores compared to all other models when tested on holdout data (Table 1), and was therefore chosen for complete case analysis and inspection (Fig. 4). The ROC curve for the imputed data was comparable to that of the complete case data, and the training LOOCV AUC was almost identical to the complete case AUC. Balanced accuracy and macro F1 score were decreased in complete case analysis compared to the training LOOCV scores but both scores were still greater than 0.7.

**Figure 4.**
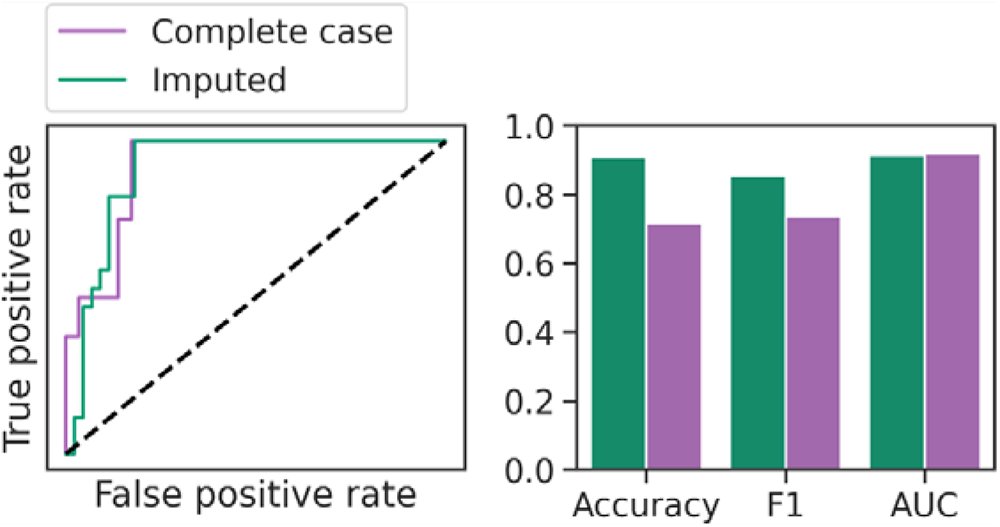
Complete case analysis for an Extra Random Forest model tasked with predicting 90 day mortality in sepsis. Performance is documented by a receiver-operating characteristic (ROC) curve (left) and a bar plot (right) showing balanced accuracy, macro F1 score, and ROC area-under-curve (AUC) score. The dotted diagonal line accompanying the ROC curves represents a model with a random performance level.

**Table 1.** Holdout performance for the top-performing model selected within each classifier family. Each model is presented as the name of the classifier family, the feature selection method that generated the optimal feature set, and the number of features selected for the top-performing model. The highest ranking metrics are highlighted in bold font. Bootstrapped 95% confidence intervals are shown in square brackets, generated using 100 rounds of resampling.

|  | Balanced accuracy | Macro F1 score | Macro AUC score |
| --- | --- | --- | --- |
| *Prediction of 30 day mortality* |  |  |  |
| Logistic regression-RFE-SVM, top 10 | 0.58 [0.50–0.67] | 0.56 [0.49–0.60] | 0.49 [0.44–0.58] |
| Linear SVM-RFE-SVM, top 10 | 0.59 [0.5–0.67] | 0.57 [0.49–0.60] | 0.52 [0.42–0.61] |
| SVM (cubic polynomial)-RFE-SVM, top10 | **0.63 [0.54–0.71]** | 0.62 [0.53–0.66] | 0.59 [0.53–0.64] |
| KNN-Boruta, top 7 | 0.63 [0.63–0.67] | **0.65 [0.64–0.71]** | 0.68 [0.64–0.72] |
| Naive Bayes-RFE-SVM, top 9 | 0.50 [0.38–0.54] | 0.50 [0.38–0.53] | 0.52 [0.44–0.57] |
| Random Forest-Boruta, top 6 | 0.46 [0.46–0.50] | 0.41 [0.40–0.42] | 0.59 [0.55–0.64] |
| Extra Random Forest-Boruta, top 7 | 0.54 [0.42–0.58] | 0.54 [0.40–0.58] | **0.73 [0.64–0.83]** |
| XGBoost-Boruta, top 7 | 0.50 [0.38–0.54] | 0.50 [0.38–0.53] | 0.48 [0.31–0.56] |
| *Prediction of 90 day mortality* |  |  |  |
| Logistic regression-RFE-SVM, top 10 | 0.71 [0.63–0.79] | 0.73 [0.64–0.79] | 0.69 [0.58–0.78] |
| Linear SVM-RFE-SVM-top 10 | 0.71 [0.63–0.79] | 0.73 [0.64–0.79] | 0.67 [0.58–0.78] |
| SVM (quartic polynomial)-RFE-SVM, top 8 | 0.70 [0.63–0.79] | 0.72 [0.64–0.79] | 0.72 [0.67–0.77] |
| KNN-RFE-SVM, top 6 | 0.54 [0.42–0.58] | 0.54 [0.40–0.58] | 0.54 [0.44–0.64] |
| Naive Bayes-RFE-SVM, top 10 | 0.71 [0.63–0.79] | 0.73 [0.64–0.79] | 0.71 [0.61–0.94] |
| Random Forest-Boruta, top 3 | 0.67 [0.58–0.75] | 0.67 [0.58–0.72] | 0.80 [0.77–0.89] |
| Extra Random Forest-RFE-SVM, top 7 | **0.75 [0.67–0.83]** | **0.79 [0.71–0.88]** | **0.85 [0.81–0.86]** |
| XGBoost-Boruta, top 6 | 0.54 [0.42–0.58] | 0.54 [0.40–0.58] | 0.58 [0.44–0.72] |
| *Prediction of Gram-negative infection* |  |  |  |
| Logistic regression-RFE-SVM, top 6 | **0.83 [0.75–1.0]** | **0.86 [0.80–1.0]** | 0.76 [0.64–1.0] |
| Linear SVM-RFE-SVM, top 5 | 0.83 [0.75–1.0] | 0.86 [0.80–1.0] | 0.71 [0.57–1.0] |
| SVM (cubic polynomial)-RFE-SVM, top 5 | 0.76 [0.68–0.93] | 0.76 [0.68–0.86] | 0.71 [0.57–0.92] |
| KNN-RFE-SVM, top 5 | 0.60 [0.43–0.68] | 0.59 [0.4–0.68] | 0.81 [0.71–0.93] |
| Naive Bayes-RFE-SVM, top 6 | 0.66 [0.50–0.75] | 0.67 [0.44–0.80] | 0.81 [0.71–1.0] |
| Random Forest-Boruta, top 6 | 0.67 [0.50–0.75] | 0.68 [0.44–0.80] | **0.86 [0.79–0.94]** |
| Extra Random Forest-Boruta, top 6 | 0.69 [0.61–0.86] | 0.67 [0.59–0.75] | 0.62 [0.50–0.79] |
| XGBoost-Boruta, top 5 | 0.60 [0.43–0.68] | 0.60 [0.4–0.68] | 0.76 [0.71–0.89] |

The proportion of T cells (as percentage of total PBMCs) was the most noteworthy feature of the Extra Random Forest model. Lower values for T cells influenced a prediction of 90 day mortality, as shown by the gradient for T cells on the beeswarm plot (Fig. 5). Excluding the percentage of T cells, the next most impactful features were elevated blood glucose, higher CXCR3 expression on CD4^+^ T cells, and lower levels of arachidonic acid (a 20-carbon chain polyunsaturated omega-6 fatty acid; C20:4). The percentage of T cells was the dominant factor in the Extra Random Forest model, but where SHAP values were only moderately high, the influence of blood glucose, CD4^+^ T cell CXCR3 expression and arachidonic acid encouraged the prediction of survival. The remaining features in the Extra Random Forest model – magnesium plasma concentration, APACHE II score, and CD25 expression on MAIT cells – appeared important for individual patients, rather than for the wider training cohort.

**Figure 5.**
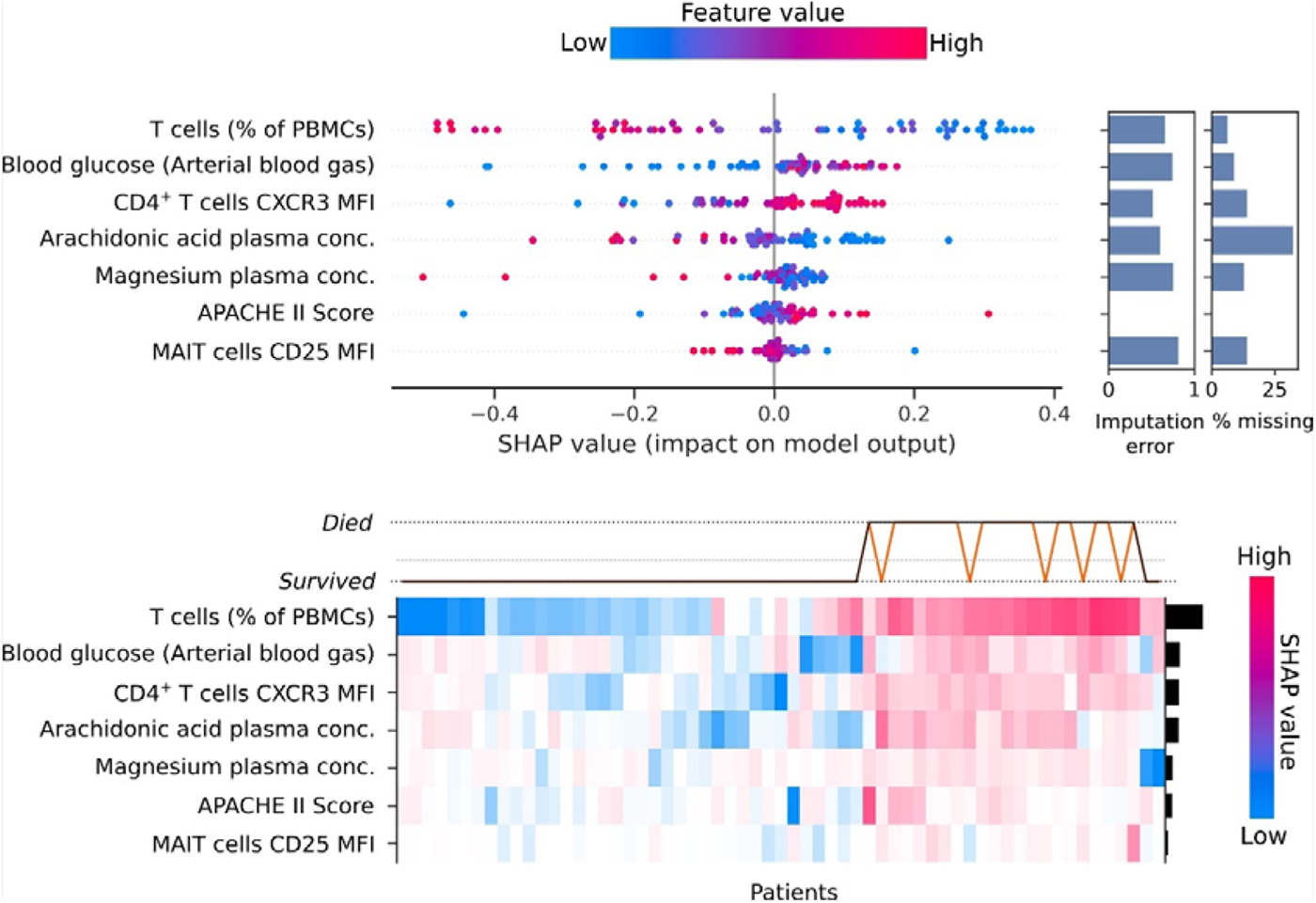
SHAP (SHapely Additive exPlanations) values for an Extra Random Forest model to predict 90 day mortality. The beeswarm plot (top) shows each observation as a single data point coloured by the value of the feature for that instance, and ranked from the most impactful on the model outcome to the least impactful. The x-axis shows the SHAP value, with a lower value corresponding to an instance having a more significant impact on the negative case for the model (*i.e.* prediction of survival), and a positive value corresponding to having a more significant impact on the positive case for the model (*i.e.* prediction of death). The bar plot on the right-hand side of the beeswarm plot shows the imputation error (with a maximum value of 1) and the percentage of missing values observed in the original data. The heatmap (bottom) shows the SHAP values for each patient. The bar plot on the right-hand y axis shows each feature’s mean absolute SHAP value as a measure of a feature’s impact on model prediction. The line plot above the heatmap displays each patient’s predicted outcome (black line) and the actual outcome (orange line). The dotted line between the possible outcomes is the expected value, equivalent to the observed mortality. Note that predictions reflect performance on the complete training data and do not reflect how the model would perform when exposed to new data.

### Neutrophils, CD8^+^ T cells and unconventional T cells form a predictive signature that differentiates Gram-negative and Gram-positive infections

With regard to the top-performing models for predicting Gram-negative cause in sepsis, logistic regression, SVMs and the Extra Random Forest model demonstrated the best LOOCV performance (Supplementary Fig. S13). The logistic regression and linear SVM models performed well on holdout data but the Random Forest model presented the best ROC AUC score overall (Table 1). The Random Forest model selected features of T cells and the neutrophil count, with the proportion of Vδ2^+^ T cells representing the feature with the highest absolute mean SHAP value (Fig. 6). The relationship between the proportion of Vδ2^+^ T cells and their SHAP values was unclear on the beeswarm plot, and was better visualised as a scatterplot (Fig. 7). As the proportion of Vδ2^+^ T cells increased, the SHAP value decreased, and thus the impact on the prediction of Gram-positive cause was stronger. Strikingly, there were two Gram-negative cases with high Vδ2^+^ γδ T cell proportions. The model successfully identified the relatively abnormal relationship these outliers had with the proportion of Vδ2^+^ T cells, and this was reflected in their low absolute SHAP values. Overall, the model displayed a strong ability to generalise despite such outliers, which highlights the added value of measuring the proportion of Vδ2^+^ T cells in the blood of sepsis patients in combination with other immunological features.

**Figure 6.**
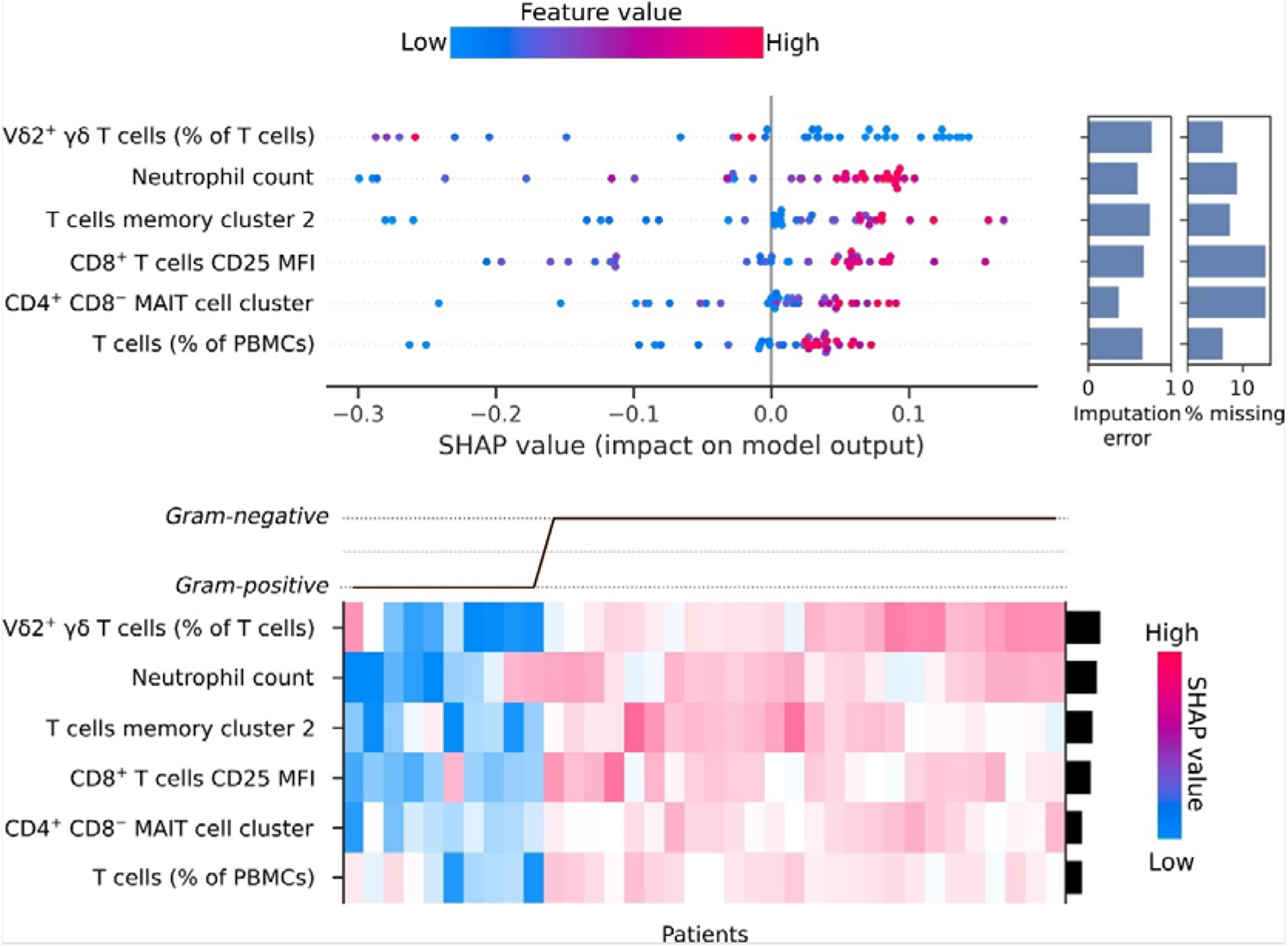
SHAP (SHapely Additive exPlanations) values for a Random Forest model to predict Gram-negative infection. A lower SHAP value corresponds to an instance having a more significant impact on the negative case for the model (*i.e.* prediction of Gram-positive sepsis), and a positive value corresponds to having a more significant impact on the positive case for the model (*i.e.* prediction of Gram-negative sepsis). The dotted line between the possible outcomes in the heatmap is the expected value, equivalent to the observed incidence of Gram-negative sepsis.

**Figure 7.**
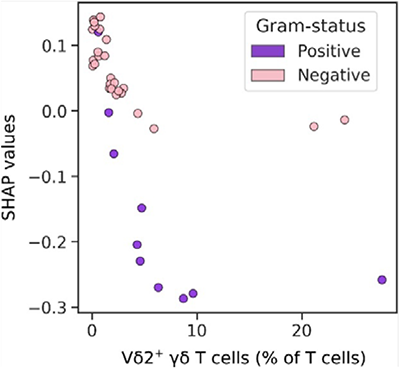
Proportion of Vδ2^+^ T cells plotted against corresponding SHAP (SHapely Additive exPlanations) values that explain the impact on a Random Forest model to predict Gram-negative infection. Each data point represents a unique patient, coloured by the causative pathogen of their acute infection.

The additional features in the Random Forest model included the total neutrophil count, CD4^+^ T cell memory cluster 2 (a CD4^+^ T cell cluster characterised by low expression of CD27 and CCR7, moderate expression of CD45RA and high expression of CD57), CD25 expression on CD8^+^ T cells, a distinct CD4^+^ CD8*^−^* MAIT cell cluster, and the proportion of T cells (as a percentage of PBMCs). Increased values for all features were associated with higher SHAP values, influencing the model to predict a Gram-negative causative pathogen, with the combination of the chosen features ultimately yielding the correct prediction. In striking contrast to these T cell related features, none of the clinical, soluble protein and lipid parameters determined for each patient were selected in the final Random Forest model, underscoring the overall importance of conventional and unconventional T cell responses for predictive models.

## DISCUSSION

We here created supervised machine learning models to predict mortality and underlying cause of infection in patients presenting with acute sepsis. A modelling pipeline was developed that considered the small cohort size, class imbalance and missing data, with a particular focus on interpretability, and employing multiple feature selection algorithms with a diverse choice of hyperparameters. Out of a total of 63 variables derived from routine clinical data, only inspired oxygen (FiO2) at the time of diagnosis differed between survivors and non-survivors after correcting for multiple comparisons; no other clinically available data were particularly informative, including previously studied biomarkers such as CRP and arterial lactate. This lack of suitable biomarkers amongst routine clinical data highlighted the need for detailed immune phenotyping to yield informative biomarkers of the patient’s response to infection.

Investigation of immune cell populations in whole blood confirmed previously well-described observations, such as decreased HLA-DR expression on monocytes [63] and a reduced proportion of circulating T cells [64, 65] amongst non-survivors. Other interesting trends were observed for soluble biomarkers such as cytokines, chemokines and acute phase protein levels in plasma. CXCL10 was decreased in those patients who died within 30 days, increased levels of IL-6 and IL-15 showed a trend towards higher odds of mortality at 30 days, and ferritin levels were higher in Gram-positive infections compared to Gram-negative infections. Ultimately, however, limited data, class imbalance and the detection limits of the assays used made it difficult to reconcile these findings.

A model for predicting 30 day mortality could not be obtained but an Extra Random Forest model for predicting 90 day mortality was identified with a holdout AUC score of 0.85, representing a considerable improvement on the majority of previously reported prognosis biomarkers [4, 13]. The 90 day mortality model showed a diverse selection of input features, including parameters that quantified immune populations, activation profiles of T cells, lipid plasma concentrations and the APACHE II severity score. The diversity of the chosen features highlights the benefits of capturing variables that describe multiple systems and how their combination can contribute to the model performance.

The proportion of T cells (as a percentage of PBMCs) was the main contributing feature to decision-making in the Extra Random Forest model. A comparison of T cells showed a significant difference between survivors and non-survivors at 90 days, in agreement with lymphopenia being a well-documented sign of increased severity and associated with higher mortality [15]. Additional features in the Extra Random Forest model included CXCR3 expression on CD4^+^ T cells, plasma concentrations of arachidonic acid and blood glucose. While the CXCR3 ligand CXCL10 was shown before to correlate with severity in sepsis [67], in the present study concentrations of CXCL10 were lower in non-survivors within 30 days. The relevance of blood glucose levels is supported by the surviving sepsis campaign international guidelines, which recommend tight control of blood glucose levels, with hyperglycaemia associated with increased mortality [18]. In addition, a reduction in arachidonic acid metabolism has been described in sepsis patients compared to healthy controls [68], and eicosanoid lipid mediators that are derived from arachidonic acid have been implicated in the pathogenesis of sepsis [69].

A compelling finding from this study was the importance attributed to unconventional T cells, even amongst the diversity of available features to select from, which included clinical variables, protein and lipid mediators as well as the proportions and phenotypes of T cells, monocytes and neutrophils in blood. In predicting 90 day mortality, increased expression of the activation marker CD25 on MAIT cells influenced survival prediction, in line with a recent description of highly activated MAIT cells in clinical and experimental sepsis [70]. The reduction in circulating Vδ2^+^ T cells and MAIT cells in patients with Gram-negative sepsis seen in the present study might reflect recruitment of these cells to sites of infection [71, 72, 73] and agrees with previous observations in septic patients [74, 75, 76] and volunteers challenged with *E. coli* [77]. In contrast, our own previous analysis in a different sepsis cohort showed increased proportions of circulating Vγ9^+^ γδ T cells in patients infected with HMB-PP^+^ pathogens [78], and others described higher levels of peripheral MAIT cells but not of γδ T cells in critically ill patients infected with *Streptococcus* spp. (*i.e*. bacteria incapable of producing the corresponding ligands), compared to non-streptococcal causes [79]. Such conflicting findings may be due to key differences in patient characteristics, time points, severity and microbiological definition as well as the methodology used for the flow cytometric analysis. Regardless of these discrepancies, unconventional T cells recently helped differentiate the causative pathogen in patients with acute peritonitis [30] and have been identified as critical players in multi-parameter immune signatures with implications in COVID-19 prognosis [80, 81]. The work presented here provides additional evidence that profiling of Vδ2^+^ T cells and MAIT cells makes valuable contributions to predictive models of acute infection. In support, a recent study described MAIT cell activation (as defined by expression of HLA-DR) combined with the APACHE II score as best indicator of 28 day mortality in sepsis patients [76].

Rigorous validation is a concern in biomarker discovery [82, 83], as it is essential to distinguish between the data used for evaluating a model and the data used for model development, especially the selection of biomarkers to be included. The generation of independent holdout data was the only mechanism to ensure that the ascertained model and feature set combination were not overfitting to the chosen training data, other than the generation of an entirely new patient cohort for validation. The primary limitation in the analysis of cytokines, chemokines and acute phase proteins in plasma were the detection limits, resulting in all but six analytes being below or above the detectable range for 20% or more of the tested samples – highlighting the intrinsic problem of multiplex analyses of biologically distinct mediators. It is also important to note that the selection of features in machine learning models and their associated SHAP values do not imply causation but rather that their combined interaction simply identifies a correlation with the predicted target. To identify the underlying cause, additional experimentation and analysis would be required. Nevertheless, machine learning models can assist in narrowing the list of target variables for investigation in subsequent experiments, thus representing a useful hypothesis-generating exercise [36]. Additionally, the interpretability of machine learning models offers the potential to identify a more general sepsis model by identifying dysregulation patterns across multiple interconnected systems, with the potential to gather insights that move beyond static dogmas of ‘cytokine storm’ and ‘immune paralysis’ [84].

A critical constraint in this study was the patient heterogeneity, reflecting the complex and poorly defined nature of sepsis pathology [85]. Fewer than 70% of the patients enrolled in this study had a microbiologically confirmed infection, a rate comparable to previous observations in sepsis [22]. It was impossible to identify whether this was due to a failure of microbiological culture or the genuine absence of any bacterial infection. Additionally, around 25% of patients were admitted to the ICU with trauma or following emergency surgery. Although this was included as a categorical variable in the machine learning pipeline to account for a potentially confounding effect, the clinical condition and type of care for such patients would differ from those that had not experienced trauma. There were also insufficient data regarding patient co-morbidity and history of infectious disease before admission to the ICU. Such data form important confounding variables for both prediction of survival and the underlying cause of infection.

There is a solid case to be made that the current definition of sepsis is inadequate, which draws focus to the dysregulated host response as the characterising feature of sepsis. In fact, most clinical trials that sought to restore the immune balance have either failed to show benefit or have proven harmful [86]. The complicated patterns of clinical presentation represent a barrier to the advancement of diagnosis and therapy, and it is increasingly being recognised that the Sepsis-3 definition cannot distinguish the complex heterogeneity observed in the pathophysiology of sepsis [87]. Research into COVID-19, a condition that has many parallels to sepsis, has reported success in uncovering immunological signatures associated with poor outcomes with links back to the underlying biological mechanisms [80]. Thus, focusing on a well-defined pathology within sepsis is likely to yield findings more readily associated with the underlying mechanism driving the immune response. Any future study expanding on the work discussed here should carefully consider the exact inclusion and exclusion criteria. Reflecting on the success of immunophenotyping of COVID-19, simple strategies could be employed to limit recruitment to those of comparable aetiology, such as culture-positive urosepsis or acute lower respiratory infection. Alternatively, a robust recruitment approach might leverage unsupervised clustering and the identification of endotypes that could be treated as distinct yet overlapping groups [88]. In the future, sepsis is likely to be recognised not as an all-embracing syndrome but rather as a group of related conditions, each characterised by specific cellular alterations and associated biomarkers.

## ACKNOWLEDGEMENTS

We are grateful to all patients and their advocates for participating in this study and to the clinicians and nurses for their cooperation, in particular Jade Cole and Helen Hill. We would also like to thank Mat Clement, Alexander Greenshields-Watson, Ann Kift-Morgan and Kristin Ladell for their help and advice throughout this study.

## ETHICS APPROVAL

Recruitment of sepsis patients was approved by the Health and Care Research Wales Research Ethics Committee under reference 17/WA/0253, protocol number SPON1609-17 and IRAS project ID 231993, and conducted according to the principles expressed in the Declaration of Helsinki.

## PATIENT CONSENT

All study participants provided written informed consent for the collection of samples and their subsequent analysis. A waiver of consent system was used when patients were unable to provide prospective informed consent due to the nature of their critical illness or therapeutic sedation at the time of recruitment. In all cases, retrospective informed consent was sought as soon as the patient recovered and regained capacity. In cases where a patient passed away before regaining capacity, the initial consultee’s approval would stand.

## DATA AVAILABILITY

The data underlying this article will be shared on reasonable request to the corresponding author.

## AUTHOR CONTRIBUTORS

Study design: R.J.B., M.P.M., A.A., M.E. Laboratory experimentation and data acquisition: R.J.B., L.R., L.M.M., S.E.B., D.A.W. Analysis and interpretation of data: R.J.B., S.M.C., B.M., A.A., M.E. Supervision: V.B.O., P.G., M.P.M., A.A., M.E. Access to clinical samples: M.P.M. Drafting of the article: R.J.B., M.E. All authors critically revised the manuscript and approved the final version to be published.

