## Supplemental information for "Conventional and unconventional T cell responses contribute to the prediction of clinical outcome and causative bacterial pathogen in sepsis patients"

**SUPPLEMENTARY INFORMATION**

**Supplementary Table S1. Comparison of survivors and non-survivors at 30 days after diagnosis of sepsis.** Continuous values are reported as the median [range] and *p* values reported using two-tailed Mann-Whitney U tests; *p* values for proportions were generated using Fisher’s exact tests.

|  | Survivors (*n=*60) | Non-survivors (*n=*17) | *p* value |
| --- | --- | --- | --- |
| Age (years) | 65.5 [18–86] | 71.0 [47–84] | 0.149 |
| Male (%) | 51.7% | 70.6% | 0.268 |
| BMI | 28.7 [17.3–51.6] | 29.4 [22.4–52.2] | 0.751 |
| APACHE II Score | 17.0 [0–30.0] | 19.0 [0–33] | 0.294 |
| Days in critical care | 9.0 [0.8–64.8] | 9.4 [1.5–34.4] | 0.980 |
| Mechanically ventilated (%) | 53.3% | 76.4% | 0.103 |
| Renal Rt (%) | 35.0% | 47.1% | 0.404 |
| Trauma/Emergency surgery | 23.3% | 23.5% | 1.000 |
| Microbiology conﬁrmed | 68.3% | 64.7% | 0.776 |

**Supplementary Table S2. Comparison of survivors and non-survivors at 90 days after diagnosis of sepsis.** Continuous values are reported as the median [range] and *p* values reported using two-tailed Mann-Whitney U tests; *p* values for proportions were generated using Fisher’s exact tests.

|  | Survivors (*n=*56) | Non-survivors (*n=*21) | *p* value |
| --- | --- | --- | --- |
| Age (years) | 65.0 [18–86] | 71.0 [47–84] | 0.067 |
| Male (%) | 51.8% | 66.7% | 0.307 |
| BMI | 28.7 [17.3–51.6] | 29.4 [22.4–52.2] | 0.496 |
| APACHE II Score | 17.0 [0–30.0] | 19.0 [0–33.0] | 0.132 |
| Days in critical care | 7.83 [0.8–64.8] | 9.9 [1.5–55.1] | 0.403 |
| Mechanically ventilated (%) | 51.8% | 76.2% | 0.070 |
| Renal Rt (%) | 32.1% | 47.6% | 0.120 |
| Trauma/Emergency surgery | 23.2% | 23.8% | 1.000 |
| Microbiology conﬁrmed | 67.9% | 66.7% | 1.000 |

**Supplementary Table S3. Comparison of Gram-negative, Gram-positive and unknown causative pathogen in sepsis patients.** Continuous values are reported as the median [range] and *p* values generated using Kruskal-Wallis H tests for independent samples; *p* values for comparison of categorical variables were generated using Fisher’s exact test.

|  | Gram-negative (*n=*33) | Gram-positive (*n=*13) | Culture–negative (*n=*25) | *p* value |
| --- | --- | --- | --- | --- |
| Age (years) | 71.0 [36.0–86.0] | 69.0 [31.0–84.0] | 60.0 [18.0–80.0] | 0.108 |
| Male (%) | 57.6% | 76.9% | 48.0% | 0.259 |
| BMI | 29.4 [20.2–52.2] | 27.8 [17.3–34.2] | 29.9 [20.2–42.7] | 0.393 |
| APACHE II Score | 19.0 [4.0–33.0] | 19.0 [8.0–25.0] | 17.0 [0.0–30.0] | 0.482 |
| Days in critical care | 7.8 [0.8–48.0] | 12.2 [1.0–64.8] | 8.8 [1.1–19.9] | 0.362 |
| Mechanically ventilated (%) | 54.5% | 53.8% | 60.0% | 0.904 |
| Renal Rt (%) | 36.4% | 61.5% | 28.0% | 0.120 |
| Trauma/Emergency surgery | 15.2% | 23.1% | 32.0 | 0.320 |
| Mortality (at 30 days) | 24.2% | 15.4% | 24.0 % | 0.720 |
| Mortality (at 90 days) | 27.3% | 23.1% | 28.0 | 0.945 |
| *Infection site (%)* |  |  |  |  |
| Abdominal | 18.2% | 23.1% |  | 0.698 |
| Respiratory | 33.3% | 38.5% |  | 0.744 |
| Urinary | 39.4% | 0% |  |  |
| Soft tissue | 3.0% | 23.1% |  | 0.062 |
| Cardiovascular | 3.0% | 0% |  |  |
| Unknown | 3.0% | 15.4% |  | 0.189 |

**Supplementary Table S4. Description of routine clinical data available for patients diagnosed with sepsis.** The total number of patients with available data is shown (*n*) along with the average value and interquartile range (IQR).

| Variable | Category | *n* | Average [IQR] |
| --- | --- | --- | --- |
| APTT | Coagulation screen | 72 | 32.61 [27.39–33.89] |
| Alanine transaminase | Liver function test | 71 | 112.5 [18.0–70.0] |
| Albumin | Bone proﬁle | 72 | 23.23 [17.0–28.0] |
| Alkaline phosphatase | Bone proﬁle | 72 | 119.03 [66.0–139.0] |
| Amylase | Amylase | 20 | 88.58 [24.25–92.88] |
| Base excess | Blood Gas Venous | 10 | 2.1 [1.02–2.82] |
| Base excess of extracellular fluid | Blood Gas Arterial | 38 | 2.36 [0.9–2.9] |
| Basophil count | Full blood count | 69 | 0.06 [0.0–0.1] |
| Bilirubin | Liver function test | 68 | 26.37 [9.0–29.5] |
| Bilirubin | Blood Gas Arterial | 13 | 19.5 [2.33–22.0] |
| C-reactive protein (CRP) | C-reactive protein | 71 | 215.27 [131.0–289.25] |
| Calcium | Bone proﬁle | 64 | 2.06 [1.93–2.17] |
| Calcium (adjusted) | Bone proﬁle | 68 | 2.31 [2.19–2.4] |
| Calcium (ionised) | Blood Gas Arterial | 69 | 1.12 [1.08–1.19] |
| Carboxyhaemoglobin | Blood Gas Arterial | 71 | 1.04 [0.77–1.2] |
| Chloride | Blood Gas Arterial | 71 | 106.62 [103.0–109.83] |
| Clauss ﬁbrinogen level | Coagulation screen | 72 | 6.46 [4.41–7.84] |
| Creatine kinase | Creatine kinase | 7 | 952.57 [60.5–482.0] |
| Creatinine | Estimated GFR | 77 | 176.86 [67.5–192.25] |
| Eosinophil count | Full blood count | 69 | 0.16 [0.1–0.2] |
| Estimated GFR | Estimated GFR | 54 | 39.72 [21.88–55.5] |
| Free T4 | Thyroid function test | 6 | 12.88 [11.5–13.42] |
| Globulin | Bone proﬁle | 71 | 32.96 [28.0–37.5] |
| Glucose | Blood Gas Arterial | 71 | 8.88 [6.52–9.75] |
| Haematocrit (Hct) | Full blood count | 77 | 0.34 [0.28–0.38] |
| Haemoglobin (Hb) | Full blood count | 77 | 113.64 [94.0–126.0] |
| High Sensitivity Troponin I | High Sensitivity Troponin | 21 | 265.91 [13.25–164.62] |
| Inspired oxygen | Blood Gas Arterial | 68 | 38.01 [24.31–45.76] |
| International normalised ratio | International normalised ratio | 5 | 3.03 [1.6–5.0] |
| Lactate | Lactate | 18 | 2.79 [1.23–3.71] |
| Lactate | Blood Gas Arterial | 71 | 2.18 [1.18–2.82] |
| Lactate dehydrogenase | Lactate dehydrogenase | 5 | 608.3 [405.0–797.5] |
| Lymphocyte count | Full blood count | 69 | 1.0 [0.63–1.3] |
| Magnesium | Magnesium | 68 | 0.83 [0.65–0.94] |
| Mean cell haemoglobin (MCH) | Full blood count | 77 | 30.28 [28.57–32.05] |
| Mean cell volume (MCV) | Full blood count | 77 | 90.75 [86.5–94.0] |
| Methaemoglobin | Blood Gas Arterial | 71 | 1.15 [0.93–1.34] |
| Monocyte count | Full blood count | 69 | 0.98 [0.65–1.2] |
| Neutrophil count | Full blood count | 69 | 12.28 [8.0–15.9] |
| Nucleated red blood cell (NRBC) count | Full blood count | 23 | 0.05 [0.0–0.1] |
| pCO2 | Blood Gas Arterial | 69 | 5.53 [4.59–6.26] |
| pCO2 | Blood Gas Venous | 36 | 5.93 [5.07–6.35] |
| pH (arterial) | Blood Gas Arterial | 70 | 7.35 [7.29–7.42] |
| pO2 (arterial) | Blood Gas Arterial | 69 | 12.25 [10.4–13.39] |
| pO2 | Blood Gas Venous | 35 | 5.82 [4.5–6.69] |
| Phosphate | Bone proﬁle | 70 | 1.39 [0.93–1.66] |
| Platelet (PLT) count | Full blood count | 72 | 234.31 [134.0–273.54] |
| Potassium | Urea and electrolytes | 77 | 4.43 [4.0–4.7] |
| Potassium | Blood Gas Arterial | 71 | 4.3 [3.88–4.4] |
| Previous CRP | Previous CRP | 9 | 123.67 [2.0–199.0] |
| Protein | Bone proﬁle | 71 | 56.88 [49.08–64.0] |
| Prothrombin time (PT) | Coagulation screen | 72 | 16.54 [12.92–17.8] |
| Red blood cell (RBC) count | Full blood count | 72 | 3.72 [3.25–4.1] |
| Red cell distribution width (RDW) | Full blood count | 77 | 14.71 [12.73–15.7] |
| SO2 | Blood Gas Arterial | 70 | 93.11 [91.74–96.88] |
| Sodium | Urea and electrolytes | 77 | 137.98 [134.0–140.5] |
| Sodium | Blood Gas Arterial | 71 | 136.65 [133.03–139.5] |
| Standard bicarbonate (arterial) | Blood Gas Arterial | 69 | 22.31 [19.44–25.37] |
| Temperature | Blood Gas Arterial | 69 | 37.22 [36.9–37.59] |
| Total Hb (calculated) | Blood Gas Arterial | 29 | 88.43 [82.5–93.2] |
| TSH | Thyroid function test | 6 | 0.8 [0.41–1.11] |
| Urea | Urea and electrolytes | 77 | 12.58 [6.14–14.86] |
| White blood cell (WBC) count | Full blood count | 72 | 15.16 [10.09–19.29] |

**Supplementary Table S5. Monoclonal antibody cocktails for flow cytometric analyses.**

| Antigen | Fluorochrome | Clone | Isotype | Manufacturer |
| --- | --- | --- | --- | --- |
| *Staining Panel 1* |  |  |  |  |
| CD11b | BV421 | ICRF44 | Mouse IgG1, *κ* | Biolegend |
| CD14 | PE-Cy7 | M5E2 | Mouse IgG2a, *κ* | Biolegend |
| CD15 | BV605 | W6D3 | Mouse IgG1, *κ* | Biolegend |
| CD19 | V500 | HIB19 | Mouse IgG1, *κ* | BD |
| CD40 | PE | MAB89 | Mouse IgG1, *κ* | Beckman Coulter |
| CD62L | PE-Cy5 | DREG-56 | Mouse IgG1, *κ* | BD |
| CD64 | APC-H7 | 10.1 | Mouse IgG1, *κ* | BD |
| CD86 | FITC | 2331 | Mouse IgG1, *κ* | Biolegend |
| HLA-DR | BV711 | L243 | Mouse IgG2a, *κ* | Biolegend |
| *Staining Panel 2* |  |  |  |  |
| CD3 | APC/FIRE | SK7 | Mouse IgG1, *κ* | Biolegend |
| CD4 | PE-Cy5.5 | S3.5 | Mouse IgG2a, *κ* | Life Technologies |
| CD8a | BV711 | RPA-T8 | Mouse IgG1, *κ* | Biolegend |
| CD14 | V500 | M5E2 | Mouse IgG2a, *κ* | BD |
| CD27 | PE-Cy7 | M-T271 | Mouse IgG1, *κ* | Biolegend |
| CD45RA | PE Dazzle | HI100 | Mouse IgG2b, *κ* | Biolegend |
| CD57 | FITC | NK-1 | Mouse IgM, *κ* | BD |
| CD161 | APC | 191B8 | Mouse IgG2a, *κ* | Miltenyi |
| CD197 (CCR7) | BV421 | G043H7 | Mouse IgG2a, *κ* | Biolegend |
| TCR-Vα7.2 | BV605 | 3C10 | Mouse IgG1, *κ* | Biolegend |
| TCR-Vδ2 | PE | B6 | Mouse IgG1, *κ* | BD |
| TCR-pan-γδ | PE-Cy5 | IMMU510 | Mouse IgG1, *κ* | Beckman Coulter |
| *Staining Panel 3* |  |  |  |  |
| CD3 | APC/FIRE | SK7 | Mouse IgG1, *κ* | Biolegend |
| CD4 | PE-Cy5.5 | S3.5 | Mouse IgG2a, *κ* | Life Technologies |
| CD8a | BV711 | RPA-T8 | Mouse IgG1, *κ* | Biolegend |
| CD14 | V500 | M5E2 | Mouse IgG2a, *κ* | BD |
| CD19 | V500 | HIB19 | Mouse IgG1, *κ* | BD |
| CD25 | PE-Cy7 | M-A251 | Mouse IgG1, *κ* | Biolegend |
| CD69 | PE-CF594 | FN50 | Mouse IgG1, *κ* | BD |
| CD161 | APC | 191B8 | Mouse IgG2a, *κ* | Miltenyi |
| CXCR3 | FITC | 49801 | Mouse IgG1, *κ* | R&D Systems |
| HLA-DR | BV421 | G46-6 | Mouse IgG2a, *κ* | BD |
| TCR-Vα7.2 | BV605 | 3C10 | Mouse IgG1, *κ* | Biolegend |
| TCR-Vδ2 | PE | B6 | Mouse IgG1, *κ* | BD |
| TCR-pan-γδ | PE-Cy5 | IMMU510 | Mouse IgG1, *κ* | Beckman Coulter |

**Supplementary Table S6.** Description of all variables considered as potential features for machine learning models. Where a variable was later removed, a reason for exclusion is given. NMAR: not missing at random.

**Supplementary Table S6. Description of all variables considered as potential features for machine learning models.** Where a variable was later removed, a reason for exclusion is given. NMAR, not missing at random; OOB, out-of-bag.

| Feature name | Category | Reason for exclusion |
| --- | --- | --- |
| Age (Years) | Demographics |  |
| Gender | Demographics |  |
| Body Mass Index (BMI) | Physiology |  |
| APACHE-II Score | Severity score |  |
| Days ventilated | Interventions |  |
| Renal Replacement Therapy (RTT) | Interventions |  |
| Emergency/Trauma | Medical history |  |
| Actual bicarbonate (aHCO3) | Blood gas analysis (Arterial) |  |
| Actual bicarbonate (aHCO3) | Blood gas analysis | *n*=1 only |
| ADAMSTS13 protease assay | Clinical laboratory measurement – ADAMST | *n*=1 only |
| Alanine Transaminase (ALT) | Clinical laboratory measurement – Liver function tests | Imputation OOB error ≥1.0 |
| Albumin | Clinical laboratory measurement – Liver function tests |  |
| Alkaline Phosphatase | Clinical laboratory measurement – Liver function tests |  |
| Amikacin | Clinical laboratory measurement – Amikacin | NMAR;  *n*=1 only |
| Ammonia | Clinical laboratory measurement – Ammonia | *n*=1 only |
| Amylase | Clinical laboratory measurement – Amylase | >40% of patients with missing data |
| Activated partial thromboplastin clotting time | Clinical laboratory measurement – Coagulation screen |  |
| Aspartate transaminase (AST) | Clinical laboratory measurement – Aspartate transaminase | *n*=2 only |
| Base excess of extracellular fluid | Blood gas analysis (Arterial) | >40% of patients with missing data |
| Base excess of extracellular fluid | Blood gas analysis (Venous) | Imputation OOB error ≥1.0 |
| Base excess | Blood gas analysis (Venous) | Imputation OOB error ≥1.0 |
| Basophil count | Clinical laboratory measurement – Full blood count |  |
| Bicarbonate | Clinical laboratory measurement – Bicarbonate |  |
| Bilirubin | Blood gas analysis (Arterial) | Imputation OOB error ≥1.0 |
| Bilirubin | Blood gas analysis (Venous) | *n*=7 only; overlaps with liver functional tests |
| Bilirubin | Clinical laboratory measurement – Liver function tests |  |
| C-reactive protein | Clinical laboratory measurement – C-reactive protein |  |
| Albumin adjusted calcium | Clinical laboratory measurement – Calcium |  |
| Albumin adjusted calcium | Clinical laboratory measurement – Liver function tests | Merged with calcium request |
| Ionised Calcium | Blood gas analysis (Arterial) |  |
| Ionised Calcium | Blood gas analysis (Venous) | >40% of patients with missing data |
| Calcium | Clinical laboratory measurement – Calcium |  |
| Carboxyhaemoglobin | Blood gas analysis (Arterial) |  |
| Carboxyhaemoglobin | Blood gas analysis (Venous) | >40% of patients with missing data |
| Chloride | Blood gas analysis (Arterial) |  |
| Chloride | Blood gas analysis (Venous) | >40% of patients with missing data |
| Chloride | Clinical laboratory measurement – Chloride | *n*=1 only; overlaps with blood gas analysis |
| Cholesterol | Clinical laboratory measurement – Cholesterol |  |
| Clauss fibrinogen level | Clinical laboratory measurement – Coagulation screen |  |
| Cortisol | Clinical laboratory measurement – Cortisol | NMAR;  *n*=1 only |
| Creatine kinase | Clinical laboratory measurement – Creatine Kinase | *n*=7 only |
| Creatinine ratio (Urine) | Clinical laboratory measurement – Protein | *n*=1 only |
| Creatinine (Urine) | Clinical laboratory measurement – Protein | *n*=1 only |
| Creatinine | Clinical laboratory measurement – Electrolyte profile |  |
| Creatinine | Clinical laboratory measurement – Estimated GFR | Merged with urea and electrolytes request |
| Creatinine | Clinical laboratory measurement – Urea and electrolytes | Merged with urea and electrolytes request |
| Digoxin | Clinical laboratory measurement – Digoxin | NMAR;  *n*=1 only |
| Eosinophil count | Clinical laboratory measurement – Full blood count |  |
| Estimated GFR | Clinical laboratory measurement – Electrolyte profile | Merged with estimated GFR request |
| Estimated GFR | Clinical laboratory measurement – Estimated GFR |  |
| Factor VII level | Clinical laboratory measurement – Factor VII | *n*=2 only |
| Ferritin | Clinical laboratory measurement – Ferritin | *n*=1 only |
| Folate | Clinical laboratory measurement – Folate | *n*=1 only |
| Free T4 | Clinical laboratory measurement – Thyroid function test | *n*=2 only |
| Gamma-glutamyl transferase | Clinical laboratory measurement – Gamma-glutamyl transferase | *n*=1 only |
| Gentamicin | Clinical laboratory measurement – Gentamicin | NMAR;  *n*=5 only |
| Globulin | Clinical laboratory measurement – Globulin |  |
| Glucose (Random) | Clinical laboratory measurement – Glucose (Random) | *n*=4 only |
| Glucose | Blood gas analysis (Arterial) |  |
| Glucose | Blood gas analysis (Venous) | >40% of patients with missing data |
| Haematocrit (Hct) | Clinical laboratory measurement – Full blood count |  |
| Haemoglobin (Hb) | Clinical laboratory measurement – Full blood count |  |
| Haemoglobin (Hb) | Blood gas analysis | *n*=1 only |
| HDL Cholesterol | Clinical laboratory measurement – Lipid profile | *n*=1 only |
| HDL Ratio | Clinical laboratory measurement – Lipid profile | *n*=1 only |
| High sensitivity troponin I | Clinical laboratory measurement – High sensitivity troponin I | Imputation OOB error ≥1.0 |
| Inspired oxygen | Blood gas analysis (Arterial) |  |
| Inspired oxygen | Blood gas analysis (Venous) | >40% of patients with missing data |
| Lactate dehydrogenase | Clinical laboratory measurement – Lactate dehydrogenase | *n*=5 only |
| Lactate | Clinical laboratory measurement – Lactate |  |
| Lactate | Blood gas analysis (Arterial) |  |
| Lactate | Blood gas analysis (Venous) | >40% of patients with missing data |
| LDL Cholesterol | Clinical laboratory measurement – Lipid profile | *n*=1 only |
| Lymphocyte count | Clinical laboratory measurement – Full blood count |  |
| Magnesium | Clinical laboratory measurement – Liver function tests | Merged with magnesium request |
| Magnesium | Clinical laboratory measurement – Magnesium |  |
| Mean cell haemoglobin | Clinical laboratory measurement – Full blood count |  |
| Mean cell volume | Clinical laboratory measurement – Full blood count |  |
| Methaemoglobin | Blood gas analysis | *n*=1; overlaps with arterial blood gas analysis |
| Methaemoglobin | Blood gas analysis (Arterial) |  |
| Methaemoglobin | Blood gas analysis (Venous) |  |
| Monocyte count | Clinical laboratory measurement – Full blood count |  |
| Neutrophil count | Clinical laboratory measurement – Full blood count |  |
| Non-HDL cholesterol | Clinical laboratory measurement – Lipid profile | *n*=1 only |
| Nucleated red blood cell count | Clinical laboratory measurement – Full blood count |  |
| Oxyhaemoglobin | Blood gas analysis (Arterial) | *n*=2 only |
| pCO2 | Blood gas analysis | *n*=1; overlaps with arterial blood gas analysis |
| pCO2 | Blood gas analysis (Arterial) |  |
| pCO2 | Blood gas analysis (Venous) |  |
| pH | Blood gas analysis | *n*=1; overlaps with arterial blood gas analysis |
| pH | Blood gas analysis (Arterial) |  |
| Phosphate | Clinical laboratory measurement – Liver function tests | Merged with phosphate request |
| Phosphate | Clinical laboratory measurement – Phosphate |  |
| Platelet count | Clinical laboratory measurement – Full blood count |  |
| pO2 | Blood gas analysis | *n*=1; overlaps with arterial blood gas analysis |
| pO2 | Blood gas analysis (Arterial) |  |
| pO2 | Blood gas analysis (Venous) | Imputation OOB error ≥1.0 |
| Potassium | Blood gas analysis (Arterial) |  |
| Potassium | Blood gas analysis (Venous) |  |
| Potassium | Clinical laboratory measurement – Electrolyte profile | Imputation OOB error ≥1.0 |
| Potassium | Clinical laboratory measurement – Urea and electrolytes |  |
| Procalcitonin | Clinical laboratory measurement – Procalcitonin | *n*=1 only |
| Protein (Urine) | Clinical laboratory measurement – Protein | *n*=1 only |
| Protein | Clinical laboratory measurement – Liver function tests |  |
| Prothrombin time | Clinical laboratory measurement – Coagulation screen |  |
| Red blood cell count | Clinical laboratory measurement – Full blood count |  |
| Red cell distribution width | Clinical laboratory measurement – Full blood count |  |
| Reptilase clotting time | Clinical laboratory measurement – Reptilase clotting time | *n*=1 only |
| Reticulocytes | Clinical laboratory measurement – Reticulocyte count | *n*=1 only |
| SO2 | Blood gas analysis | *n*=1; overlaps with arterial blood gas analysis |
| SO2 | Blood gas analysis (Arterial) |  |
| Sodium | Blood gas analysis (Arterial) |  |
| Sodium | Blood gas analysis (Venous) |  |
| Sodium | Clinical laboratory measurement – Electrolyte profile | Imputation OOB error ≥1.0 |
| Sodium | Clinical laboratory measurement – Urea and electrolytes |  |
| Standard bicarbonate | Blood gas analysis |  |
| Standard bicarbonate | Blood gas analysis (Arterial) |  |
| Temperature | Blood gas analysis (Arterial) |  |
| Temperature | Blood gas analysis (Venous) |  |
| Thrombin time | Clinical laboratory measurement – Thrombin time | *n*=2 only |
| Total CO2 | Blood gas analysis | *n*=1 only |
| Total Hb calculated | Blood gas analysis (Arterial) |  |
| Total Hb calculated | Blood gas analysis (Venous) | Imputation OOB error ≥1.0 |
| Triglyceride | Clinical laboratory measurement – Lipid profile | *n*=1 only |
| Triglyceride | Clinical laboratory measurement – Triglyceride | *n*=1 only |
| TSH (Thyroid-stimulating hormone) | Clinical laboratory measurement – Thyroid function tests | *n*=6 only |
| Urate | Clinical laboratory measurement – Urate | *n*=3 only |
| Urea | Clinical laboratory measurement – Urea | Merged with urea and electrolyte request |
| Urea | Clinical laboratory measurement – Liver function tests | Merged with urea and electrolyte request |
| Urea | Clinical laboratory measurement – Electrolyte profile | Merged with urea and electrolyte request |
| Urea | Clinical laboratory measurement – Urea and electrolytes |  |
| Urine volume | Clinical laboratory measurement – Creatinine clearance | *n*=1 only |
| Vancomycin | Clinical laboratory measurement – Vancomycin | NMAR;  *n*=1 only |
| Vitamin B12 | Clinical laboratory measurement – Vitamin B12 | *n*=1 only |
| White blood cell count | Clinical laboratory measurement – Full blood count |  |
| ——————————————— | ——————————————— |  |
| CCL5 plasma concentration | Luminex |  |
| CXCL10 plasma concentration | Luminex |  |
| IL-4 plasma concentration | Luminex |  |
| Lactoferrin plasma concentration | Luminex |  |
| MMP-8 plasma concentration | Luminex |  |
| MMP-9 plasma concentration | Luminex |  |
| PD-L1 plasma concentration | Luminex |  |
| VEGF plasma concentration | Luminex |  |
| IL-6 plasma concentration | ELISA |  |
| CCL2 in plasma >29.9 pg/ml | Luminex |  |
| CXCL13 in plasma >20.3 pg/ml | Luminex |  |
| FLT3L in plasma >14.7 pg/ml | Luminex |  |
| G-CSF in plasma >23.8 pg/ml | Luminex |  |
| IL-1α in plasma >4.8 pg/ml | Luminex |  |
| IL-10 in plasma >4.0 pg/ml | Luminex |  |
| IL-15 in plasma >6.3 pg/ml | Luminex |  |
| CXCL8 in plasma >3.8 pg/ml | Luminex |  |
| OSM in plasma >369.7 pg/ml | Luminex |  |
| Procalcitonin in plasma >2110 pg/ml | Luminex |  |
| Ferritin in plasma >6120 pg/ml | Luminex |  |
| IFN-γ in plasma >14.7 pg/ml | ELISA |  |
| TNFα in plasma >7.7 pg/ml | ELISA |  |
| ——————————————— | ——————————————— |  |
| C2 carnitine plasma concentration | Lipids |  |
| C3 carnitine plasma concentration | Lipids |  |
| C4 carnitine plasma concentration | Lipids |  |
| C6 carnitine plasma concentration | Lipids |  |
| C8 carnitine plasma concentration | Lipids |  |
| C10 carnitine plasma concentration | Lipids |  |
| C12 carnitine plasma concentration | Lipids |  |
| C14 carnitine plasma concentration | Lipids |  |
| C16 carnitine plasma concentration | Lipids |  |
| C18 carnitine plasma concentration | Lipids |  |
| C18:1 carnitine plasma concentration | Lipids |  |
| C12-2OH/3OH plasma concentration | Lipids |  |
| C8:0 plasma concentration | Lipids |  |
| C10:0 plasma concentration | Lipids |  |
| C12:0 plasma concentration | Lipids |  |
| C14:0 plasma concentration | Lipids |  |
| C16:0 plasma concentration | Lipids |  |
| C18:0 plasma concentration | Lipids |  |
| C18:1 plasma concentration | Lipids |  |
| C18:2 plasma concentration | Lipids |  |
| C18:3 plasma concentration | Lipids |  |
| C20:4 plasma concentration | Lipids |  |
| C20:5 plasma concentration | Lipids |  |
| C22:6 plasma concentration | Lipids |  |
| ——————————————— | ——————————————— |  |
| T cells (% of PBMCs) | Flow cytometry – Major subsets |  |
| Monocytes (% of Leukocytes) | Flow cytometry – Major subsets |  |
| Neutrophils (% of Leukocytes) | Flow cytometry – Major subsets |  |
| CD4^+^ CD8^−^ T cells (% of T cells) | Flow cytometry – T cell subsets |  |
| CD4^−^ CD8^+^ T cells (% of T cells) | Flow cytometry – T cell subsets |  |
| Vδ2^+^ γδ T cells (% of T cells) | Flow cytometry – T cell subsets |  |
| Vα7.2^+^ CD161^+^ MAIT cells (% of T cells) | Flow cytometry – T cell subsets |  |
| T cells memory cluster 0 (% of T cells) | Flow cytometry – Memory T cell clusters |  |
| T cells memory cluster 1 (% of T cells) | Flow cytometry – Memory T cell clusters |  |
| T cells memory cluster 2 (% of T cells) | Flow cytometry – Memory T cell clusters |  |
| T cells memory cluster 3 (% of T cells) | Flow cytometry – Memory T cell clusters |  |
| T cells memory cluster 4 (% of T cells) | Flow cytometry – Memory T cell clusters | Imputation OOB error ≥1.0 |
| T cells memory cluster 14 (% of T cells) | Flow cytometry – Memory T cell clusters |  |
| T cells activation cluster 0 (% of T cells) | Flow cytometry – Activated T cell clusters T cells |  |
| T cells activation cluster 1 (% of T cells) | Flow cytometry – Activated T cell clusters T cells |  |
| T cells activation cluster 4 (% of T cells) | Flow cytometry – Activated T cell clusters T cells |  |
| T cells activation cluster 6 (% of T cells) | Flow cytometry – Activated T cell clusters T cells |  |
| T cells activation cluster 7 (% of T cells) | Flow cytometry – Activated T cell clusters T cells |  |
| T cells activation cluster 8 (% of T cells) | Flow cytometry – Activated T cell clusters T cells |  |
| T cells activation cluster 10 (% of T cells) | Flow cytometry – Activated T cell clusters T cells |  |
| T cells activation cluster 12 (% of T cells) | Flow cytometry – Activated T cell clusters T cells |  |
| T cells activation cluster 13 (% of T cells) | Flow cytometry – Activated T cell clusters T cells |  |
| T cells activation cluster 14 (% of T cells) | Flow cytometry – Activated T cell clusters T cells |  |
| T cells activation cluster 16 (% of T cells) | Flow cytometry – Activated T cell clusters T cells |  |
| Vδ2^+^ memory cluster 0 (% of Vδ2^+^ T cells) | Flow cytometry – Vδ2^+^ Memory T cell clusters |  |
| Vδ2^+^ memory cluster 1 (% of Vδ2^+^ T cells) | Flow cytometry – Vδ2^+^ Memory T cell clusters |  |
| Vδ2^+^ memory cluster 2 (% of Vδ2^+^ T cells) | Flow cytometry – Vδ2^+^ Memory T cell clusters |  |
| Vδ2^+^ memory cluster 3 (% of Vδ2^+^ T cells) | Flow cytometry – Vδ2^+^ Memory T cell clusters | Imputation OOB error ≥1.0 |
| Vδ2^+^ memory cluster 4 (% of Vδ2^+^ T cells) | Flow cytometry – Vδ2^+^ Memory T cell clusters |  |
| Vδ2^+^ activation cluster 0 (% of Vδ2^+^ T cells) | Flow cytometry – Activated Vδ2^+^ T cell clusters |  |
| Vδ2^+^ activation cluster 1 (% of Vδ2^+^ T cells) | Flow cytometry – Activated Vδ2^+^ T cell clusters |  |
| Vδ2^+^ activation cluster 2 (% of Vδ2^+^ T cells) | Flow cytometry – Activated Vδ2^+^ T cell clusters |  |
| Vδ2^+^ activation cluster 3 (% of Vδ2^+^ T cells) | Flow cytometry – Activated Vδ2^+^ T cell clusters |  |
| Vδ2^+^ activation cluster 4 (% of Vδ2^+^ T cells) | Flow cytometry – Activated Vδ2^+^ T cell clusters |  |
| MAIT memory cluster 0 (% of MAIT cells) | Flow cytometry – MAIT Memory T cell clusters |  |
| MAIT memory cluster 1 (% of MAIT cells) | Flow cytometry – MAIT Memory T cell clusters |  |
| MAIT memory cluster 2 (% of MAIT cells) | Flow cytometry – MAIT Memory T cell clusters |  |
| MAIT memory cluster 3 (% of MAIT cells) | Flow cytometry – MAIT Memory T cell clusters |  |
| MAIT memory cluster 4 (% of MAIT cells) | Flow cytometry – MAIT Memory T cell clusters |  |
| MAIT memory cluster 5 (% of MAIT cells) | Flow cytometry – MAIT Memory T cell clusters |  |
| MAIT memory cluster 6 (% of MAIT cells) | Flow cytometry – MAIT Memory T cell clusters |  |
| MAIT activation cluster 0 (% of MAIT cells) | Flow cytometry – Activated MAIT T cell clusters |  |
| MAIT activation cluster 1 (% of MAIT cells) | Flow cytometry – Activated MAIT T cell clusters |  |
| MAIT activation cluster 2 (% of MAIT cells) | Flow cytometry – Activated MAIT T cell clusters |  |
| MAIT activation cluster 3 (% of MAIT cells) | Flow cytometry – Activated MAIT T cell clusters |  |
| MAIT activation cluster 4 (% of MAIT cells) | Flow cytometry – Activated MAIT T cell clusters |  |
| Monocyte cluster 0 (% of Monocytes) | Flow cytometry – Monocyte clusters |  |
| Monocyte cluster 1 (% of Monocytes) | Flow cytometry – Monocyte clusters |  |
| Monocyte cluster 2 (% of Monocytes) | Flow cytometry – Monocyte clusters |  |
| Monocyte cluster 3 (% of Monocytes) | Flow cytometry – Monocyte clusters |  |
| Monocyte cluster 4 (% of Monocytes) | Flow cytometry – Monocyte clusters |  |
| Neutrophil cluster 0 (% of Neutrophils) | Flow cytometry – Neutrophil clusters |  |
| Neutrophil cluster 1 (% of Neutrophils) | Flow cytometry – Neutrophil clusters |  |
| Neutrophil cluster 2 (% of Neutrophils) | Flow cytometry – Neutrophil clusters |  |
| Neutrophil cluster 3 (% of Neutrophils) | Flow cytometry – Neutrophil clusters |  |
| Monocytes HLA-DR MFI | Flow cytometry – Monocyte activation markers |  |
| Monocytes CD40 MFI | Flow cytometry – Monocyte activation markers |  |
| Monocytes CD62L MFI | Flow cytometry – Monocyte activation markers |  |
| Monocytes CD64 MFI | Flow cytometry – Monocyte activation markers |  |
| Monocytes CD86 MFI | Flow cytometry – Monocyte activation markers |  |
| Neutrophils HLA-DR MFI | Flow cytometry – Neutrophil activation markers |  |
| Neutrophils CD40 MFI | Flow cytometry – Neutrophil activation markers |  |
| Neutrophils CD62L MFI | Flow cytometry – Neutrophil activation markers |  |
| Neutrophils CD64 MFI | Flow cytometry – Neutrophil activation markers |  |
| Neutrophils CD86 MFI | Flow cytometry – Neutrophil activation markers |  |
| CD8^+^ T cells HLA-DR MFI | Flow cytometry – CD8^+^ T cell activation markers |  |
| CD8^+^ T cells CD25 MFI | Flow cytometry – CD8^+^ T cell activation markers |  |
| CD8^+^ T cells CD69 MFI | Flow cytometry – CD8^+^ T cell activation markers |  |
| CD8^+^ T cells CD161 MFI | Flow cytometry – CD8^+^ T cell activation markers |  |
| CD8^+^ T cells CXCR3 MFI | Flow cytometry – CD8^+^ T cell activation markers |  |
| CD4^+^ T cells HLA-DR MFI | Flow cytometry – CD4^+^ T cell activation markers |  |
| CD4^+^ T cells CD25 MFI | Flow cytometry – CD4^+^ T cell activation markers |  |
| CD4^+^ T cells CD69 MFI | Flow cytometry – CD4^+^ T cell activation markers |  |
| CD4^+^ T cells CD161 MFI | Flow cytometry – CD4^+^ T cell activation markers |  |
| CD4^+^ T cells CXCR3 MFI | Flow cytometry – CD4^+^ T cell activation markers |  |
| MAIT cells HLA-DR MFI | Flow cytometry – MAIT cell activation markers |  |
| MAIT cells CD25 MFI | Flow cytometry – MAIT cell activation markers |  |
| MAIT cells CD69 MFI | Flow cytometry – MAIT cell activation markers |  |
| MAIT cells CXCR3 MFI | Flow cytometry – MAIT cell activation markers |  |
| Vδ2^+^ T cells HLA-DR MFI | Flow cytometry – Vδ2^+^ T cell activation markers |  |
| Vδ2^+^ T cells CD25 MFI | Flow cytometry – Vδ2^+^ T cell activation markers |  |
| Vδ2^+^ T cells CD69 MFI | Flow cytometry – Vδ2^+^ T cell activation markers |  |
| Vδ2^+^ T cells CD161 MFI | Flow cytometry – Vδ2^+^ T cell activation markers |  |
| Vδ2^+^ T cells CXCR3 MFI | Flow cytometry – Vδ2^+^ T cell activation markers |  |

**Supplementary Table S7. MRM transitions, MS parameters and deuterated internal standards (same MS settings) for free fatty acid and acyl carnitine analyses.**

| Lipid  (lipid carbons: double bonds) | DP  (Volts) | CE  (Volts) | Q1  *m/z* | Q3  *m/z* | Internal std (IST)^¥^  (Concentration in IST mix, µM) | Q1  *m/z*  (IST) | Q3  *m/z*  (IST) |
| --- | --- | --- | --- | --- | --- | --- | --- |
| *Free Fatty Acid* |  |  |  |  |  |  |  |
| Acetic acid (C2:0) | -35 | -26 | 194.1 | 137 | Acetic acid-d3 (50) | 197.1 | 137 |
| Propionic acid (C3:0) | -49 | -27 | 208.1 | 137 | Propionic acid-d2 (15) | 210.1 | 137 |
| Lactic acid (C3-OH) | -76 | -26 | 224.1 | 137 | “ | “ | “ |
| Butyric acid (4:0) | -90 | -28 | 222 | 137 | Butyric acid-d3 (15) | 225.1 | 137 |
| Hexanoic acid (6:0) | -100 | -30 | 250.2 | 137 | Hexanoic acid-d3 (25) | 253.2 | 137 |
| Octanoic acid (8:0) | -120 | -34 | 278 | 137 | Octanoic acid-d15 (10) | 293.3 | 137 |
| Decanoic acid (10:0) | -127 | -36 | 306 | 137 | Decanoic acid-d3 (25) | 309.3 | 137 |
| Lauric acid (12:0) | -134 | -41 | 334 | 137 | Lauric acid-d23 (10) | 357.5 | 137 |
| C12-2OH/3OH | -115 | -36 | 350.3 | 137 | “ | “ | “ |
| Myristic acid (14:0) | -120 | -42 | 362.4 | 137 | Myristic acid-d27 (10) | 389.5 | 137 |
| Palmitic acid (16:0) | -137 | -44 | 390.4 | 137 | Palmitic acid-d2 (25) | 392.4 | 137 |
| Stearic acid (18:0) | -140 | -50 | 418.5 | 137 | Stearic acid-d35 (25) | 453.7 | 137 |
| Oleic acid (18:1) | -138 | -48 | 416.5 | 137 | “ | “ | “ |
| Linoleic acid (18:2) | -140 | -44 | 414.5 | 137 | “ | “ | “ |
| Linolenic acid (18:3) | -134 | -42 | 412.4 | 137 | “ | “ | “ |
| Arachidonic acid (20:4) | -145 | -42 | 438.5 | 137 | Arachidonic acid-d8 (25) | 446.5 | 137 |
| Eicosapentaenoic acid (20:5) | -135 | -42 | 436.5 | 137 | “ | “ | “ |
| Docosahexaenoic acid (22:6) | -148 | -44 | 462.3 | 137 | “ | “ | “ |
| *Acylcarnitines* |  |  |  |  |  |  |  |
| Acetylcarnitine (C2:0) | 76 | 27 | 339.3 | 220 | Acetylcarnitine-d3 (2.5) | 342.3 | 220 |
| Propionylcarnitine (C3:0) | 76 | 27 | 353.0 | 220 | Propionylcarnitine-d3 (2.5) | 356.0 | 220 |
| Butyrylcarnitine (C4:0) | 76 | 35 | 367.3 | 220 | Butyrylcarnitine-d3 (2.5) | 370.3 | 220 |
| Hexanoylcarnitine (C6:0) | 91 | 29 | 395.3 | 220 | Hexanoylcarnitine-d3 (2.5) | 398.3 | 220 |
| Octanoylcarnitine (C8:0) | 101 | 39 | 423.4 | 220 | Octanoylcarnitine-d3 (2.5) | 426.4 | 220 |
| Decanoylcarnitine (C10:0) | 106 | 33 | 451.4 | 220 | Decanoylcarnitine-d3 (2.5) | 454.4 | 220 |
| Lauroylcarnitine (C12:0) | 111 | 35 | 479.5 | 220 | Lauroylcarnitine-d3 (2.5) | 482.5 | 220 |
| Myristoylcarnitine (C14:0) | 128 | 36 | 507.3 | 220 | Myristoylcarnitine-d3 (2.5) | 510.3 | 220 |
| Palmitoylcarnitine (C16:0) | 129 | 37 | 535.3 | 220 | Palmitoylcarnitine-d3 (2.5) | 338.3 | 220 |
| Stearoylcarnitine (C18:0) | 128 | 39 | 563.4 | 220 | Stearoylcarnitine-d3 (2.5) | 566.4 | 220 |
| Oleoylcarnitine (C18:1) | 134 | 44 | 561.3 | 220 | “ | “ | “ |

^¥^Internal standards were sourced from Sigma-Aldrich (Poole, Dorset, UK); Supelco (via Sigma-Aldrich, Poole, Dorset, UK) and Cambridge Isotope Laboratories, Inc (Tewksbury, MA, USA).

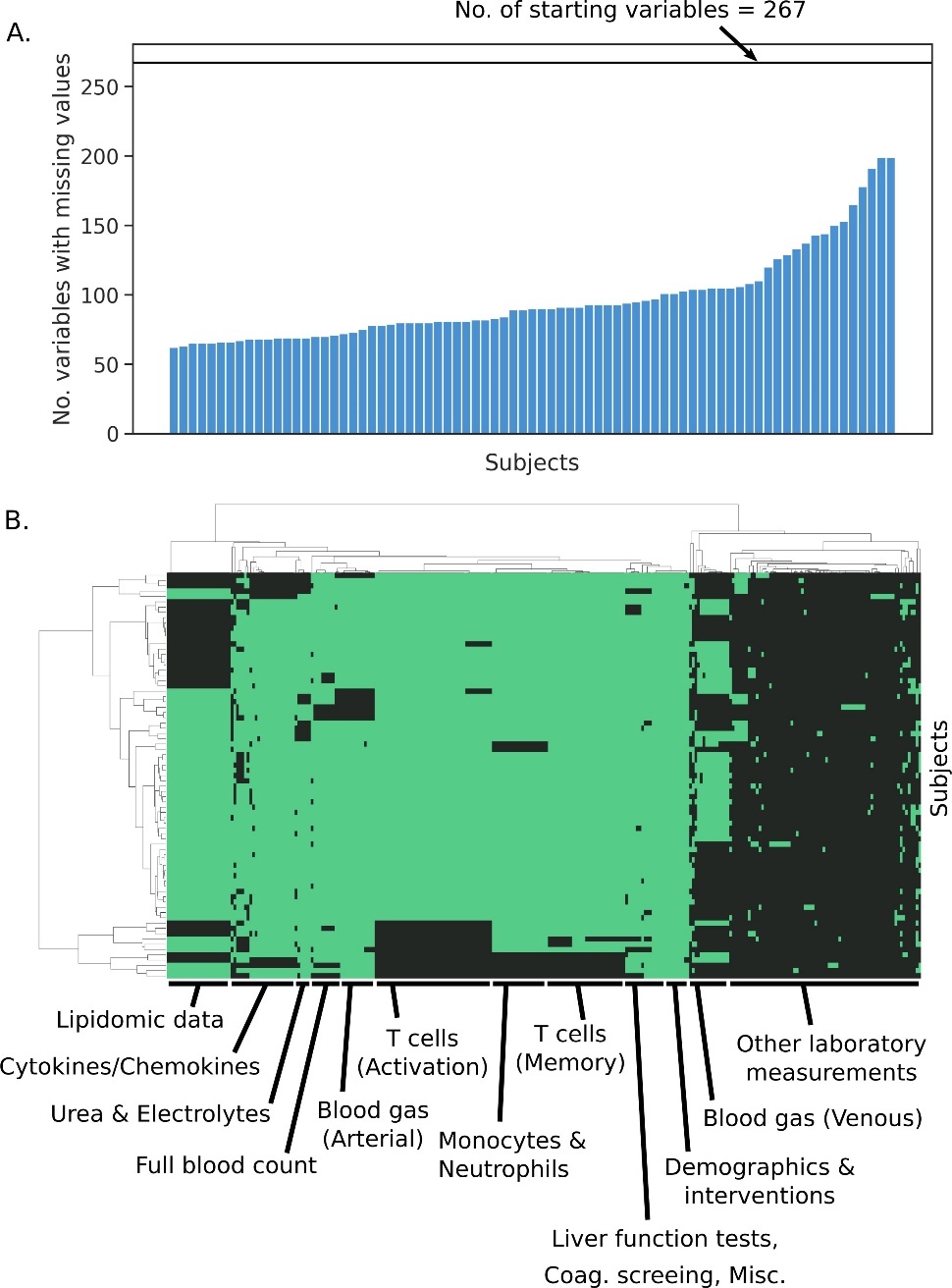

**Supplementary Figure S1. Visualisation of missing data.** (***A***) Number of variables with a value missing for each patient, out of a total of 267 available variables (shown by the black horizontal line). (***B***) Clustered heatmap with the 267 available features as columns and each patient represented by a row. A black cell represents the absence of a variable for a given patient. Rows and columns were clustered using Ward hierarchical clustering.

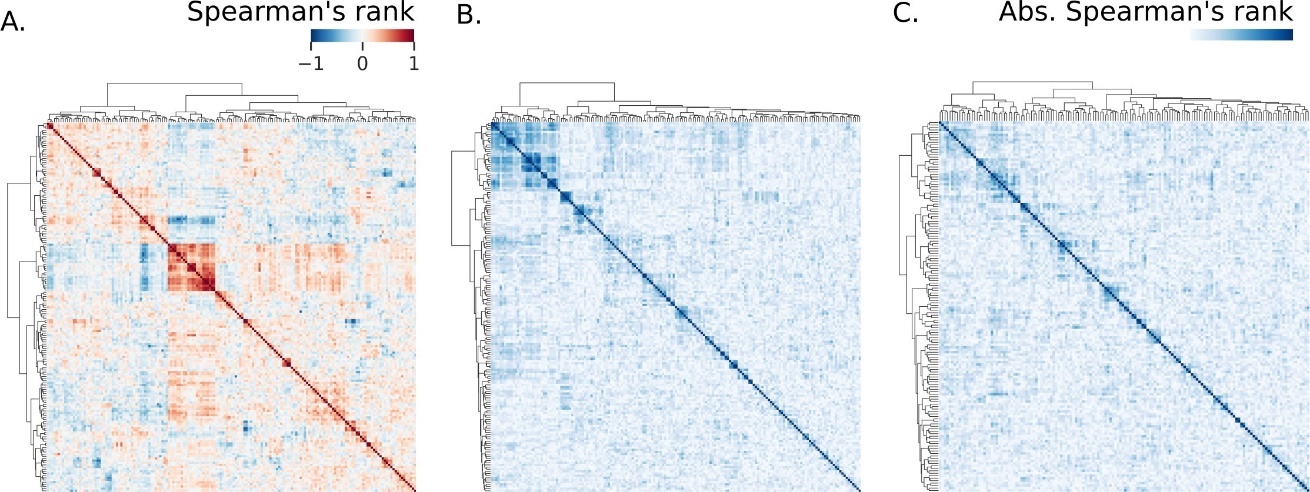

**Supplementary Figure S2. Multicollinearity amongst features visualised using pairwise Spearman’s rank correlation coefﬁcient.**  (***A***) Clustered matrix of pairwise Spearman’s rank correlation coefﬁcient shows groups of highly correlated features. The absolute pairwise Spearman’s rank correlation coefﬁcient matrix was clustered and is shown before (***B***) and after (***C***) removal of redundancies and replacement of highly correlated groups with latent variables.

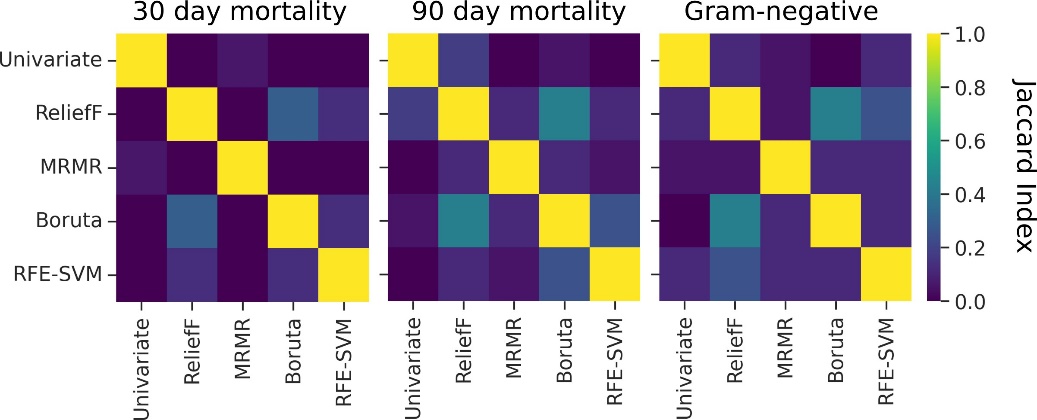

**Supplementary Figure S3. Pairwise Jaccard Index measures the overlap of feature sets generated by ﬁve independent feature selection algorithms.** Feature selection was performed for three binary target variables: mortality 30 days after diagnosis with sepsis (left), 90 days after diagnosis with sepsis (middle), and a Gram-negative causative pathogen amongst those with a positive culture (right). A Jaccard index of 1 indicated that both sets were identical; when no common features were shared, the index was 0.

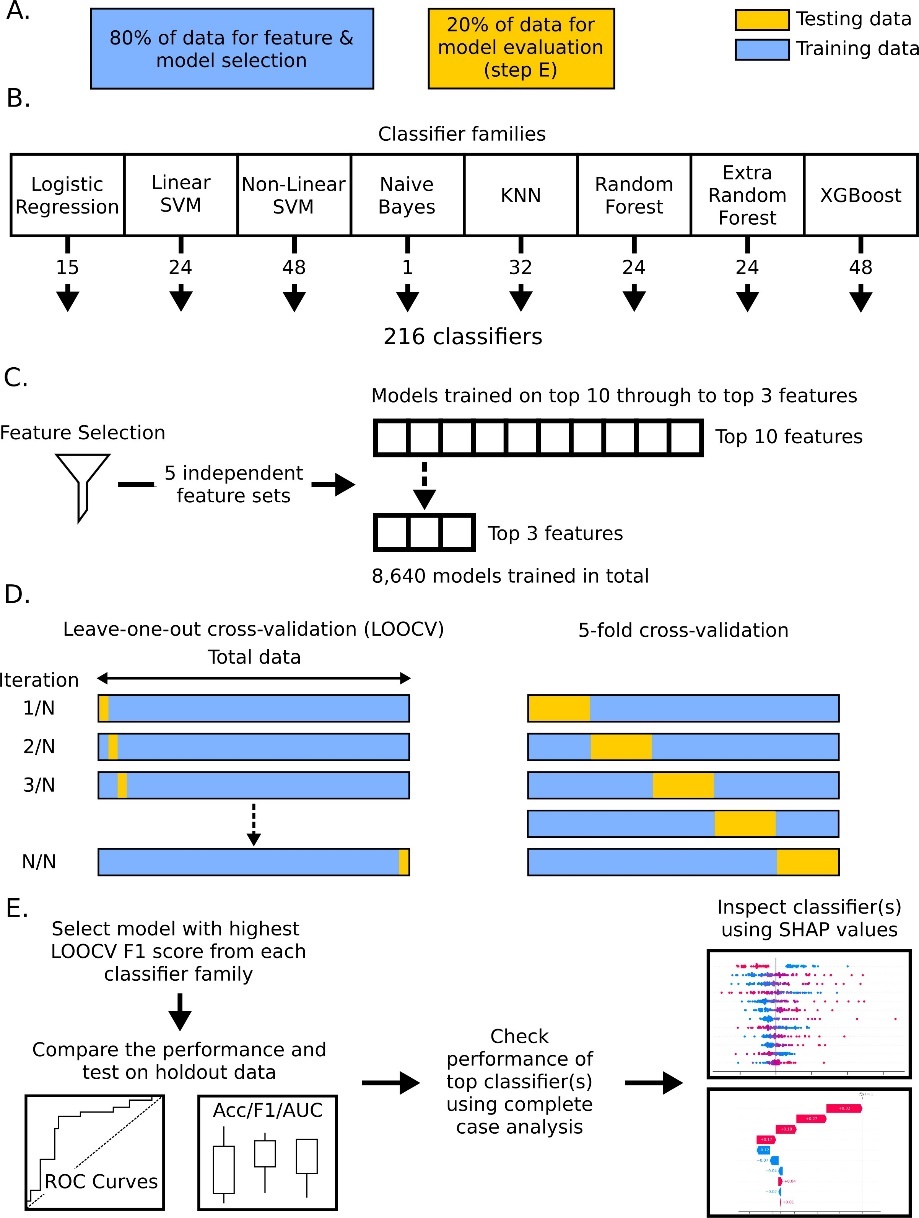

**Supplementary Figure S4. Schematic of the modelling pipeline for selecting, comparing, and inspecting classiﬁcation algorithms.** Data were first split (***A***) into training and holdout sets, retaining 20% of data for model evaluation. Data were then exposed to eight classiﬁer families (***B***), generating 216 classiﬁers in total after including multiple hyperparameters. Models were trained using ﬁve independent feature sets (***C***) iteratively on the top three through to the top ten features from each feature set, producing 40 models for each feature set and 8,880 models for each target variable. Models were trained and tested using leave-one-out and 5-fold cross-validation (***D***), and the best performing model (measured by LOOCV macro F1 score) was selected for each classiﬁer family. Cross-validation performance and performance on the holdout set were compared across classiﬁers using ROC curves, balanced accuracy, macro F1 score, and macro AUC score (***E***). The top performing classiﬁer(s) were tested against their equivalent complete case data as an additional validation step before inspection of model decisions using SHapely Additive exPlanations (SHAP) values.

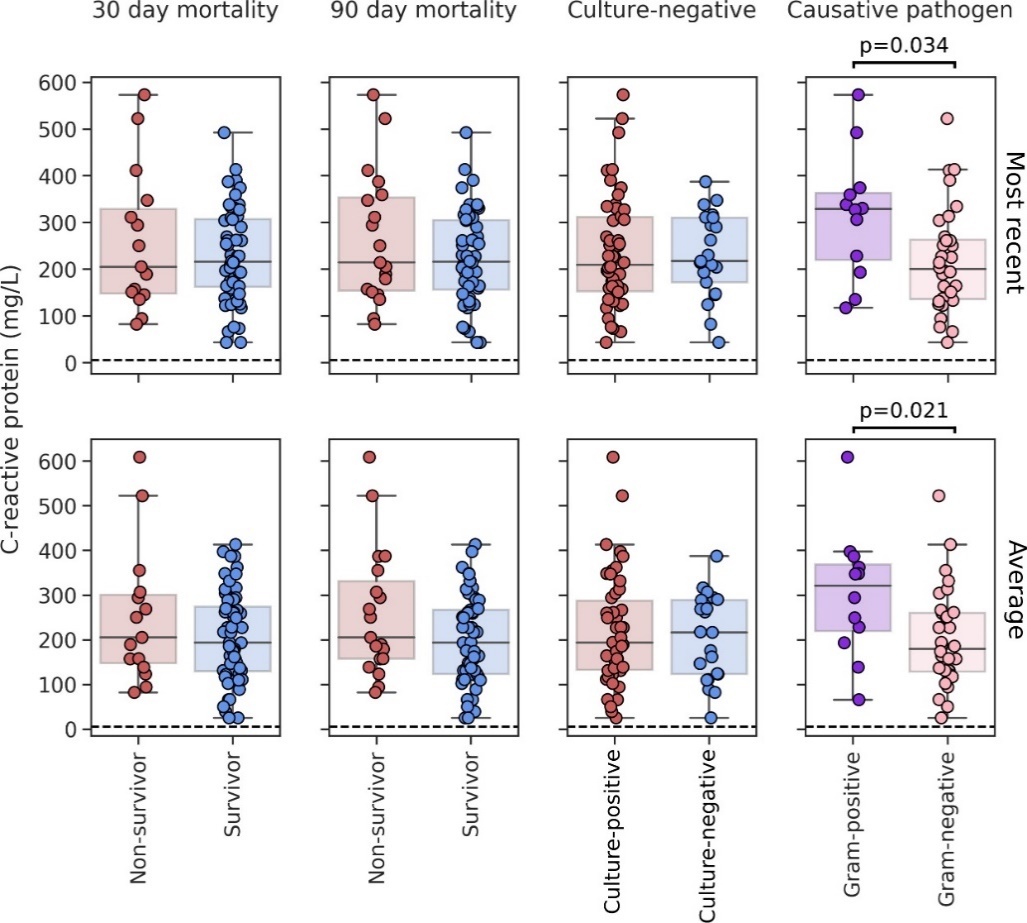

**Supplementary Figure S5. C-reactive-protein (CRP) concentration in blood of patients diagnosed with sepsis.** Values are shown for samples taken closest to enrolment time (top) and the average concentration within a window of 48 hours prior enrolment up until 8 hours after enrolment (bottom).  *p* values report comparisons using two- tailed Mann-Whitney U tests. Dotted lines represent the reference range used for CRP by Cardiff and Vale Health Board, and values above this line were considered ’raised levels of CRP’.

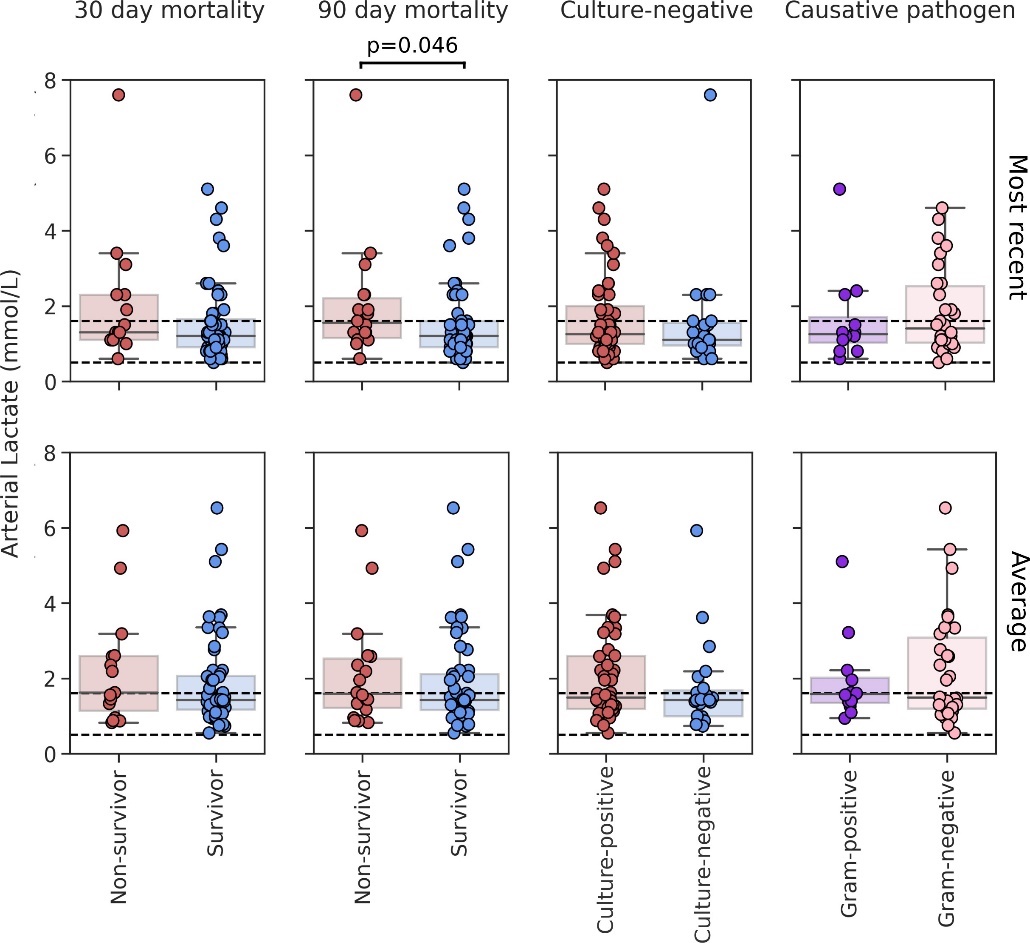

**Supplementary Figure S6. Lactate concentration in blood of patients diagnosed with sepsis.** Values are shown for samples taken closest to enrolment time (top) and the average concentration within a window of 48 hours prior enrolment up until 8 hours after enrolment (bottom). *p* values report comparison using two-tailed Mann-Whitney U tests. Dotted lines represent the reference range for blood lactate used by Cardiff and Vale Health Board, and values outside this range were considered ‘abnormal’.

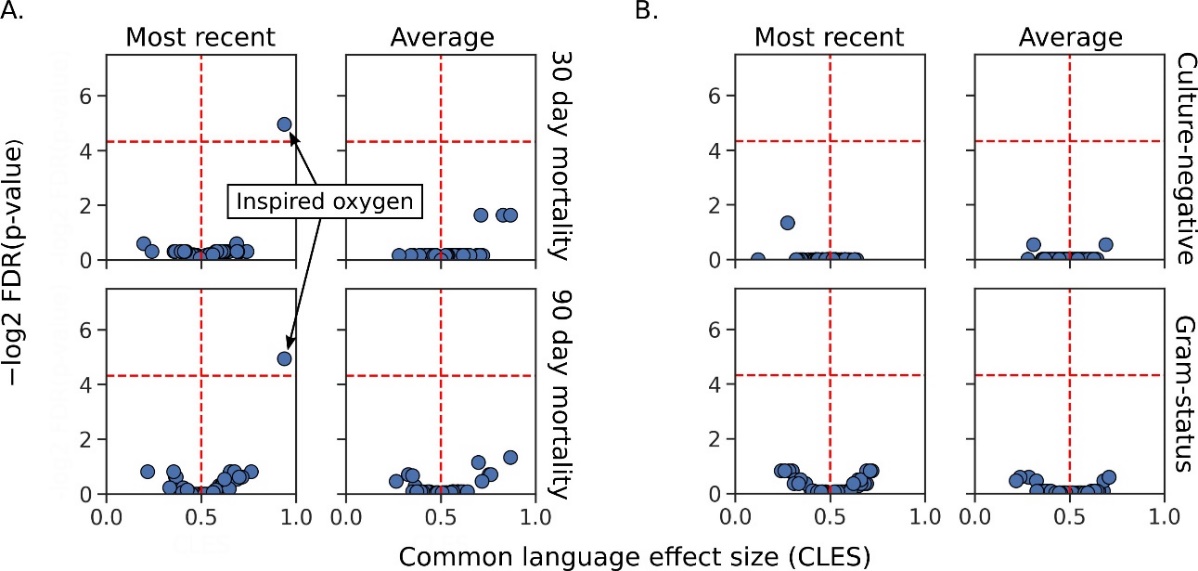

**Supplementary Figure S7. Comparisons of variables captured in routine clinical data and their ability to differentiate mortality, culture-positivity, and the Gram status of the causative pathogen in sepsis.** Data were summarised as either the most recent value relative to enrolment time or the average value within a window of 48 hours prior to enrolment and 8 hours after enrolment. (***A***) Comparisons of survivors at 30 and 90 days after enrolment to non-survivors; (***B***) differentiation based on culture results. Graphs show the common language effect size (CLES) versus the corrected *p* value for all biomarkers routinely collected under the null hypothesis that values were similar amongst patient subgroups. *p* values are reported using two-tailed non-parametric Mann-Whitney U tests with Benjamini-Hochberg corrections to control for the false discovery rate at an α of 0.05. Values below the horizontal red line have a *p* value greater than 0.05; the vertical red line represents a CLES of 50%. CLES was computed using the Pingouin package in Python and is the proportion of pairs where observations of *x* have a greater value than observations of *y*; *x* and *y* would be survivors and non-survivors, culture-negative or culture-positive sepsis, or Gram-positive and Gram-negative infections, respectively.

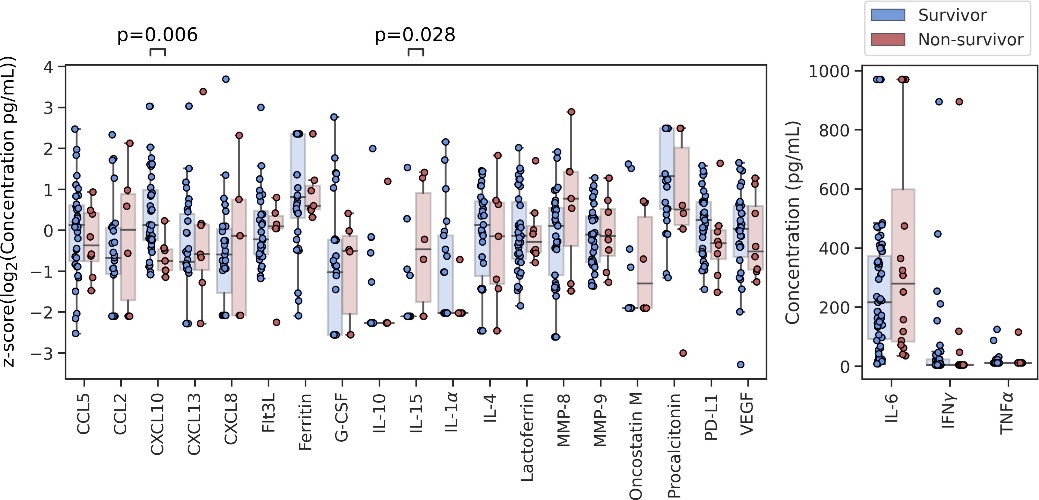

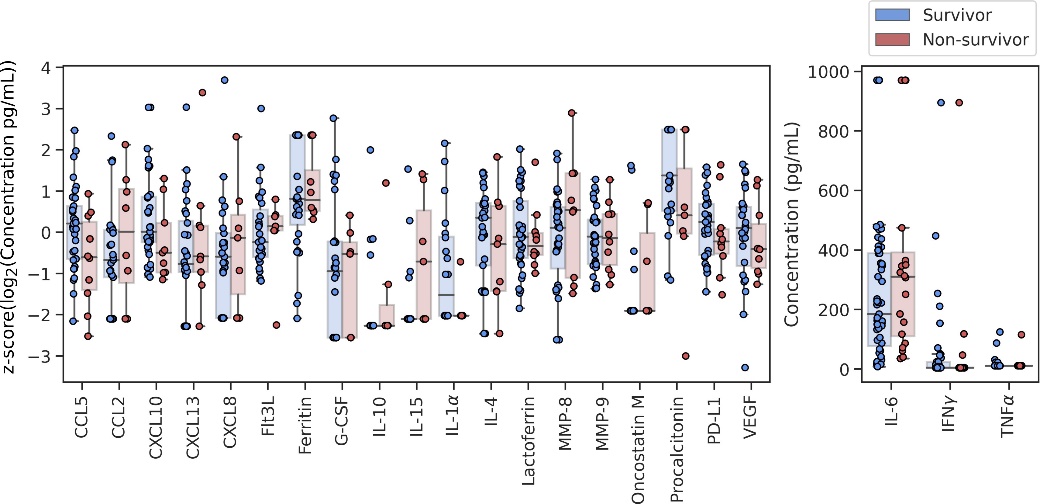

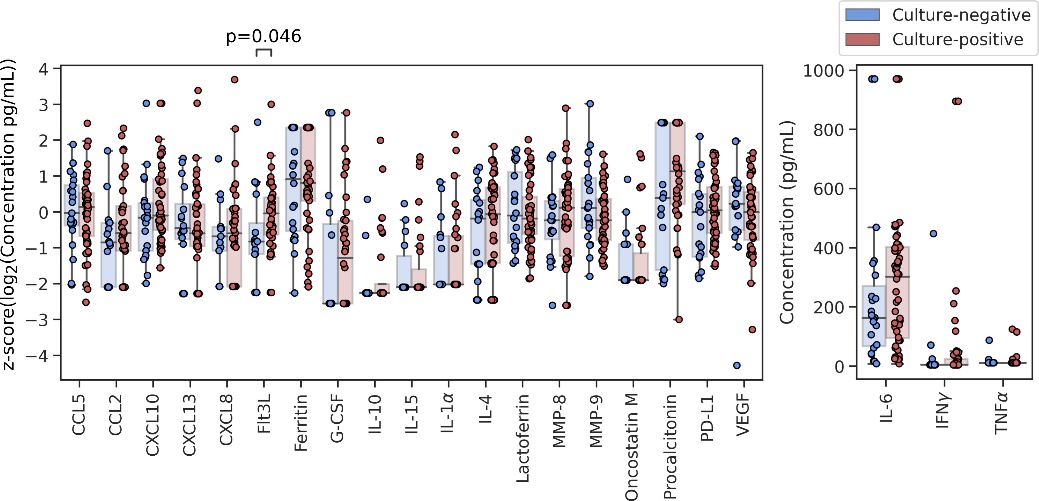

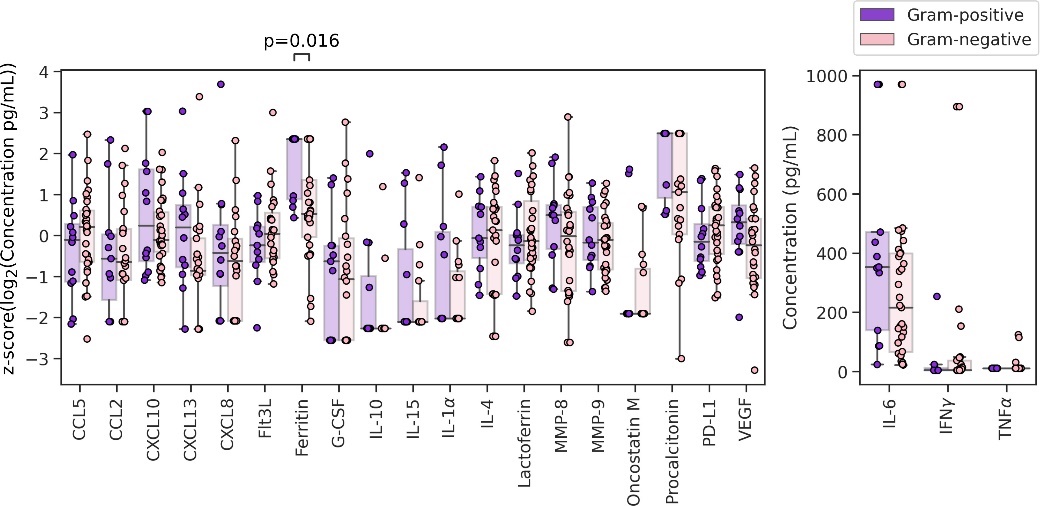

**Supplementary Figure S8. Concentration of soluble analytes in plasma of sepsis patients.** Comparisons are between survivors and non-survivors 30 days (*top panel*) and 90 days after sepsis diagnosis (*second panel*); those without and with a microbiologically conﬁrmed infection (*third panel*); and those with a Gram-positive and Gram-negative infection, amongst those with a positive bacterial culture (*bottom panel*). Signiﬁcance testing was performed using two-tailed Mann-Whitney U tests with correction for multiple comparisons made using the Benjamini-Hochberg procedure to control for false discovery rate at an α of 0.05. Some data crowded at the bottom or top of the analytical range, which reﬂected samples where concentrations of analytes were outside the detectable range of the Luminex (left) and ELISA (right) assays.

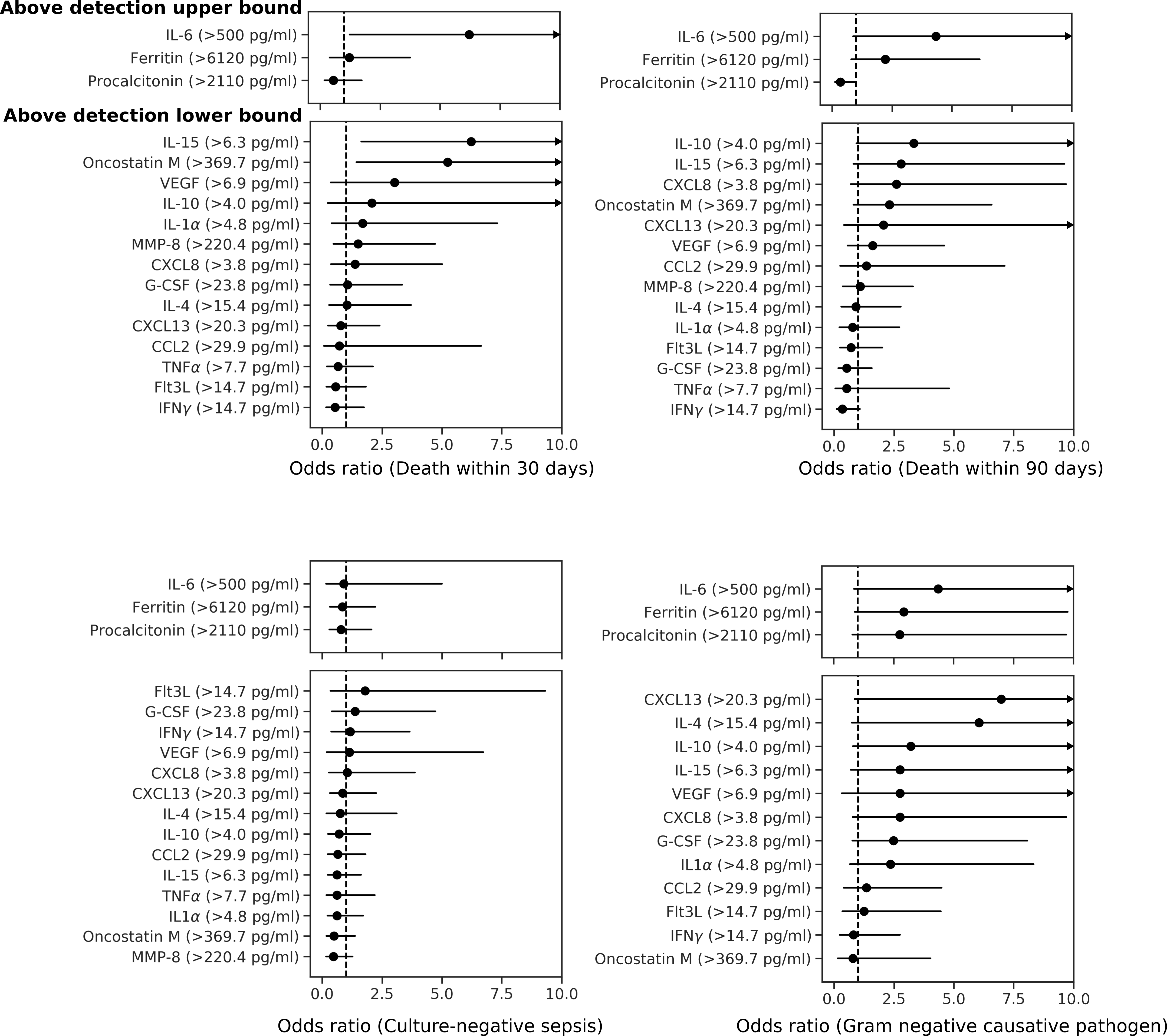

**Supplementary Figure S9. Odds ratios for death within 30 days of enrolment date (top left), death within 90 days (top right), culture-negative sepsis (bottom left), and Gram- negative infection (bottom right).** For the analytes IL-6, ferritin, and procalcitonin, patients were grouped into those above and below the upper detection limit of the assay. IL-1α, IL-4, IL-10, IL-15, MMP-8, CCL2, CXCL8, CXCL13, IFN-γ, TNF-α, G-CSF, oncostatin M, VEGF, and Flt3L were separated into those below and above the lower bound of the detection limit. CCL5, CXCL10, lactoferrin, MMP-9 and PD-L1 were excluded from this analysis because more than 90% of samples were within the detectable range. Comparisons between groups were tested for signiﬁcance using Fisher’s exact test and corrected for multiple comparisons with Benjamini-Hochberg procedure at an α of 0.05. 95% conﬁdence intervals for odds ratios were approximated as previously described by Tenny and Hoffman [49].

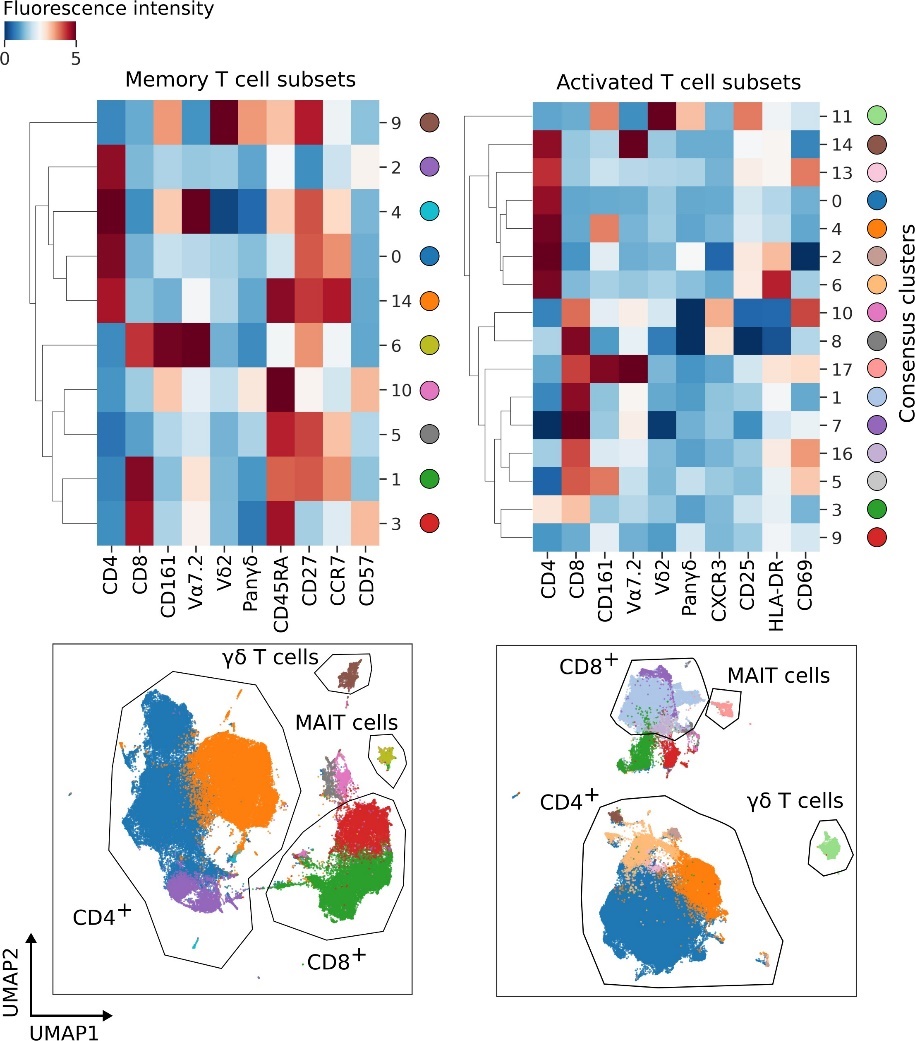

**Supplementary Figure S10. GeoWaVe ensemble clustering of T cells.** The left heatmap and UMAP scatterplot shows T cells stained for identiﬁcation of memory subsets whereas the right shows T cells stained for identiﬁcation of activated subsets. The heatmaps and accompanying UMAP scatterplots show the identiﬁed consensus clusters and their expression proﬁle. Fluorescence intensity is shown as hyperbolic arcsine with a cofactor of 150.

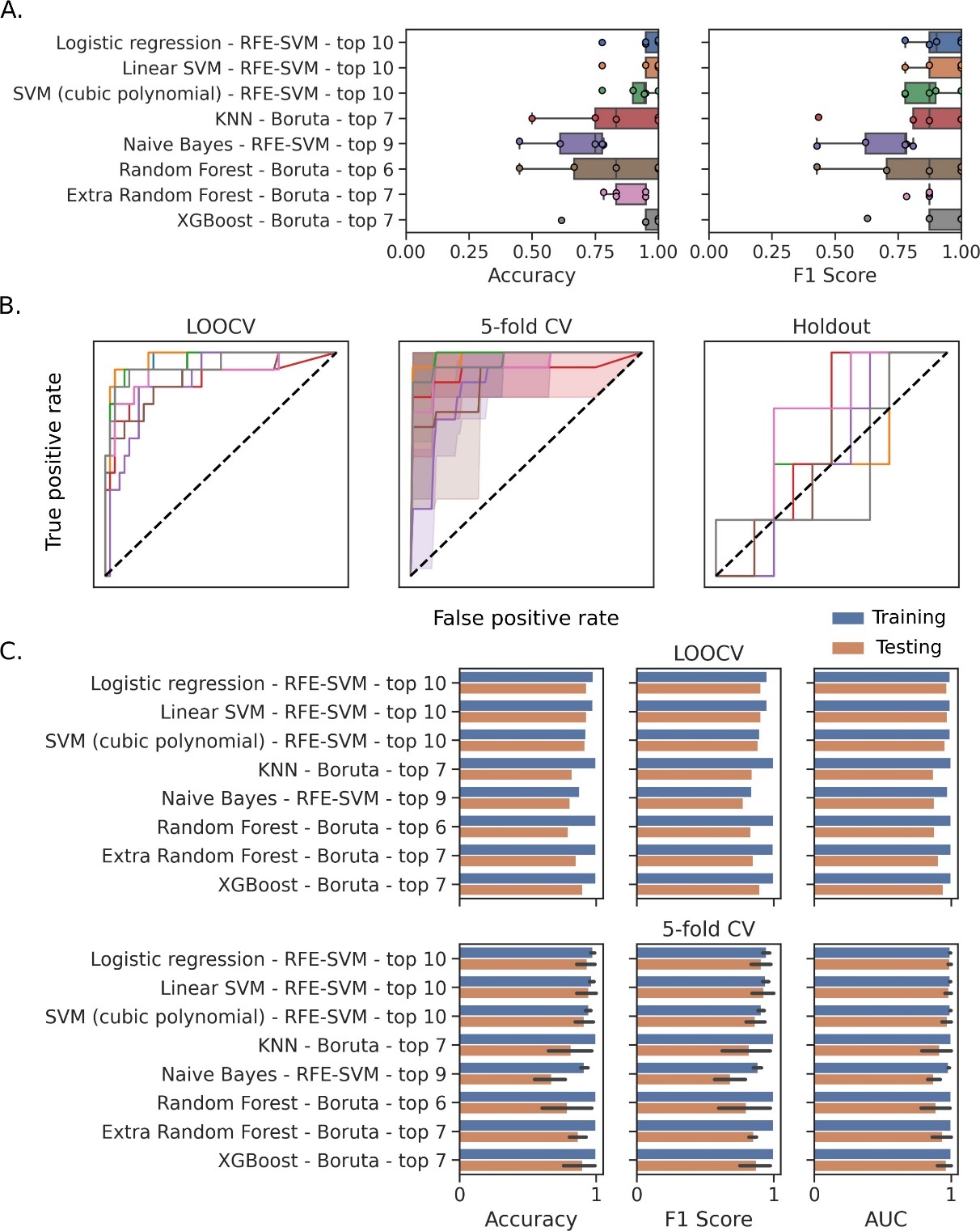

**Supplementary Figure S11. Cross-validation and holdout performance for the top-performing model selected within each classiﬁer family for predicting 30 day mortality.** Each model is presented as the name of the classiﬁer family, the feature selection method that generated the optimal feature set, and the number of features selected for the top-performing model. (***A***) Balanced accuracy and macro F1 score for each fold of 5-fold cross-validation (CV). (***B***) Model receiver-operating-characteristic (ROC) curves for leave-one-out cross-validation (LOOCV), 5-fold CV, and testing on holdout data. The dotted diagonal line represents a model with random performance level. (***C***) Difference in training and testing performance within each cross-validation procedure where error bars for 5-fold CV represent 95% bootstrap conﬁdence intervals with 1000 rounds of resampling.

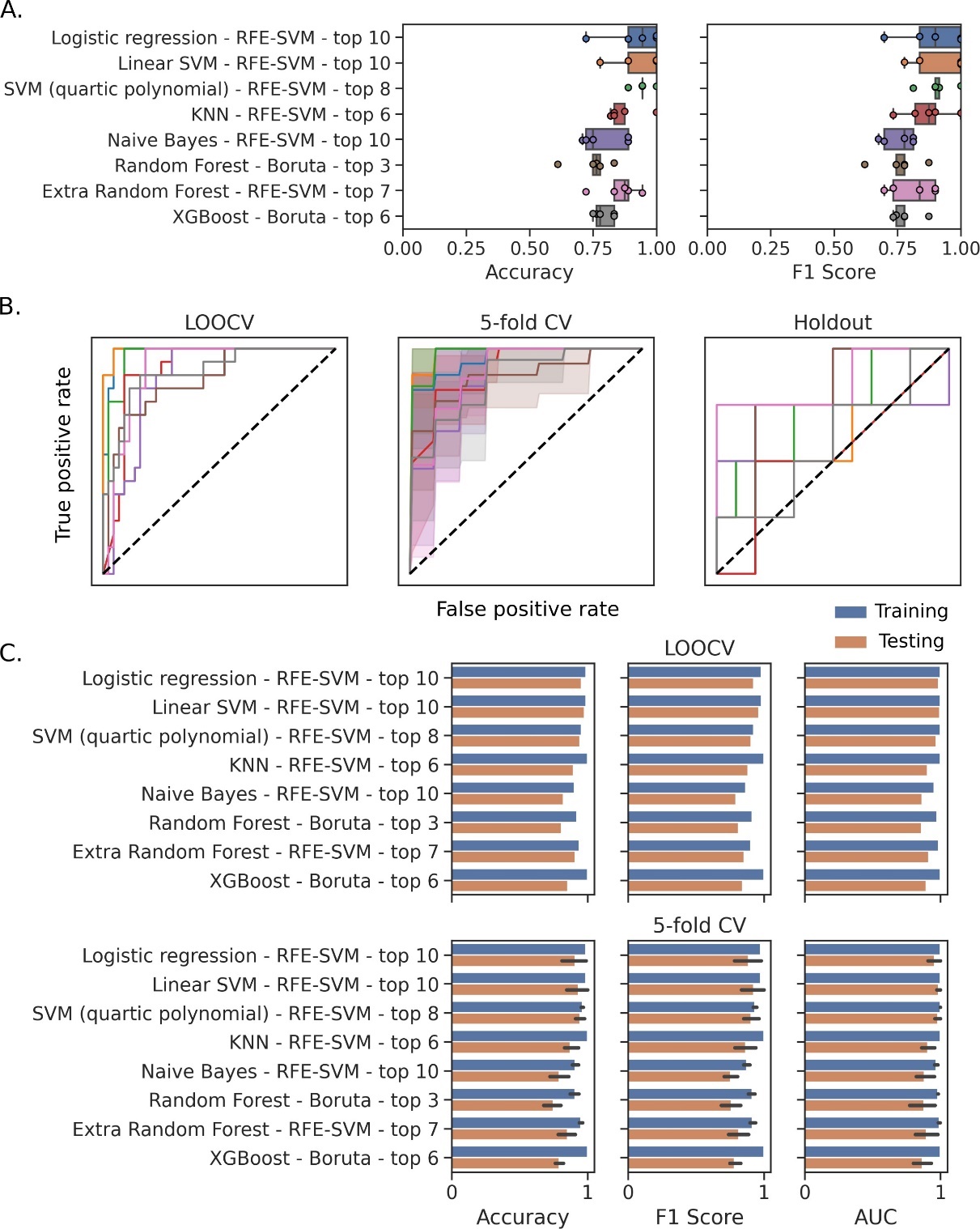

**Supplementary Figure S12. Cross-validation and holdout performance for the top-performing model selected within each classiﬁer family for predicting 90 day mortality.** Data analysis and presentation as in Supplementary Figure S11.

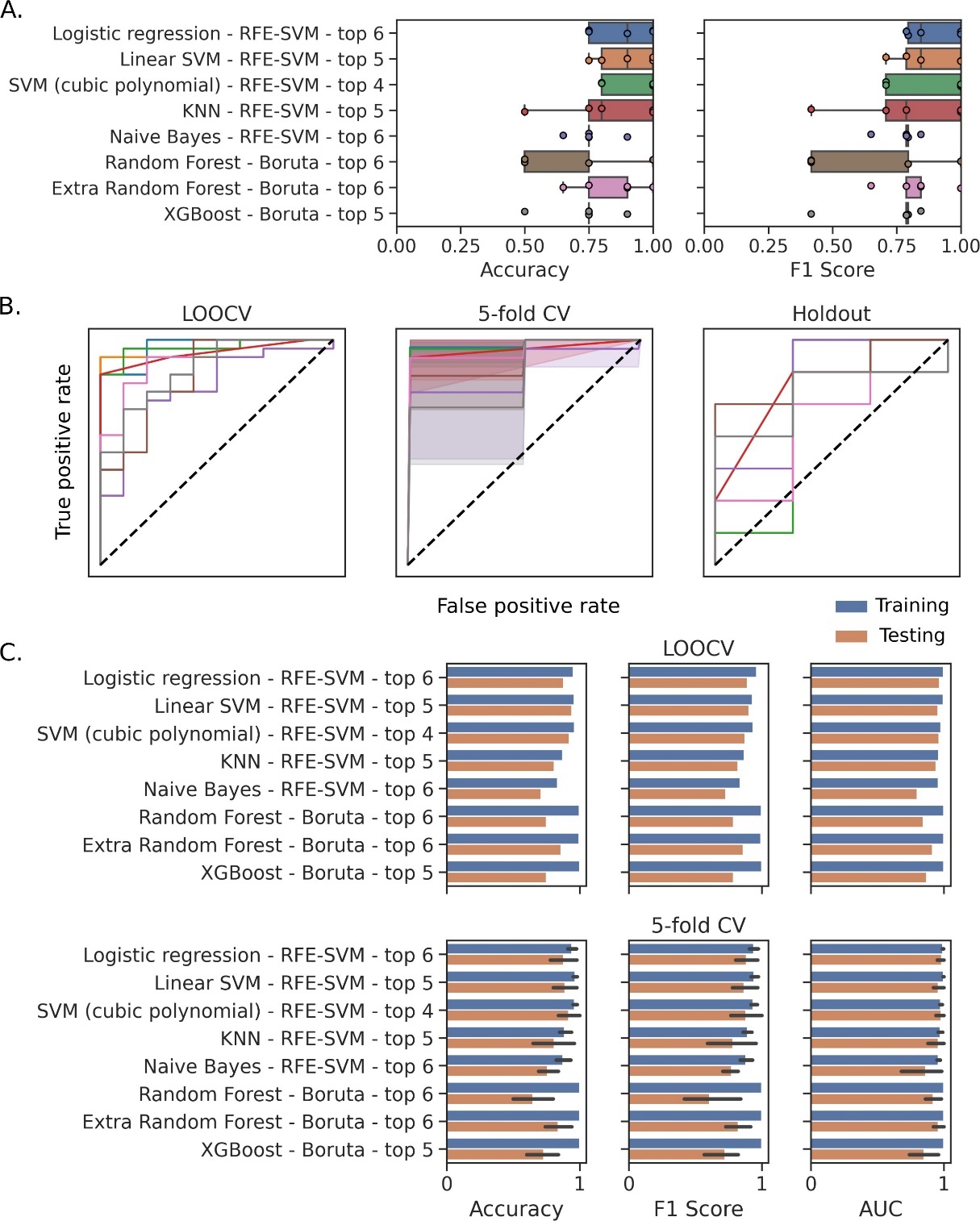

**Supplementary Figure S13. Cross-validation and holdout performance for the top-performing model selected within each classiﬁer family for predicting Gram-negative infections.** Data analysis and presentation as in Supplementary Figure S11.
